# Occupational hierarchy, racialization, and COVID-19 health outcomes among meat processing plant workers in Alberta: a community-engaged mixed-methods study

**DOI:** 10.64898/2026.05.14.26353257

**Authors:** Mohammad Yasir Essar, Eric Norrie, Edna Ramirez Cerino, Minnella Antonio, Ammar Saad, Mussie Yemane, Linda Holdbrook, Adanech Sahilie, Michael Youssef, Nour Hassan, Olivia Magwood, Samuel T. Edwards, Denise Spitzer, Annalee Coakley, Kevin Pottie, Gabriel E. Fabreau

**Author notes:** Corresponding Author: Gabriel E. Fabreau, MD, MPH, 3E28 Cal Wenzel Precision Health Building, 32380 Hospital Drive NW Calgary, Alberta, Canada T2N-4Z6, |.

## Abstract

**Background:** Meat processing plants in Alberta, Canada experienced among North America’s largest COVID-19 outbreaks. We examined health impacts among workers by occupational hierarchy and equity-relevant characteristics.

**Methods:** This exploratory sequential mixed-methods study was guided by community-based participatory research and the PROGRESS-Plus framework. Multilingual qualitative interviews and surveys using validated instruments were conducted among meat plant workers who experienced outbreaks. Interviews were analysed using inductive-deductive thematic analysis. Multivariable logistic regression and linear regression estimated associations between occupational group, racialization, facility, and self-reported COVID-19 diagnosis, physical and mental health, and mean Everyday Discrimination Scale score. We integrated findings using joint displays.

**Findings:** Qualitative and integrated analysis of thirty-six interviews described occupational hierarchy shaping unequal protection, limited communication, constrained agency, and psychosocial harms, amplified by income insecurity and family separation. Among 187 survey respondents, compared with general labour, skilled labour (aOR 0·38; 95% CI 0·15–0·89) and management (aOR 0·13; 95% CI 0·01–0·75) had lower odds of reported COVID-19 diagnosis. Compared with Black workers, other racialized workers had lower odds of reporting fair or poor mental (aOR 0·24; 95% CI 0·09–0·58) and physical health (aOR 0·20; 95% CI 0·06–0·54). Compared with workers from the primary facility, others reported lower mean everyday discrimination scores (β = –0·54; 95% CI –0·96 to –0·12).

**Interpretation:** COVID-19 harms followed workplace social hierarchies. Pandemic preparedness should combine infection-control measures with paid sick leave and income protection, multilingual communication, enforceable anti-discrimination standards, and independent reporting mechanisms.

**Funding:** Canadian Institutes for Health Research (CIHR Application no. 469206).

**Research in Context:** *Evidence before this study:* We searched PubMed/MEDLINE, Scopus, and Web of Science from June 2020 to December 2025, using terms for COVID-19, meat processing, meatpacking, occupation, and workers including migrants, racialized workers, refugees or immigrants for empirical studies published without language restrictions. Existing studies showed that meat processing plants were sites of occupational COVID-19 outbreaks and that immigrant and racialized workers experienced disproportionate infections and adverse health outcomes. The literature described pre-existing structural vulnerabilities in these settings including crowded working conditions, inadequate occupational protections, and barriers related to language, job security, and access to health information. These inequities intensified during the pandemic, leading to disproportionate infection rates, morbidity, mortality, and psychosocial stress.

*Added value of this study:* This exploratory sequential mixed-methods study used the PROGRESS-Plus framework and a community-based participatory research approach to examine COVID-19-related health impacts after large outbreaks among meat processing plant workers in Alberta, Canada. By integrating multilingual qualitative interviews with quantitative survey data, the study identified how occupational hierarchy, racialization, and processing plant shaped self-reported COVID-19 diagnosis, physical and mental health, and experiences of discrimination. The study also centres workers’ perspectives to show how workplace hierarchy, unequal communication, and limited agency contributed to health inequities during the pandemic.

*Implications of all the available evidence:* Previous and current findings suggest that COVID-19 harms in meat processing plants were shaped by pre-existing structural and workplace inequities rather than by exposure alone. Working conditions in large meat processing plants were already difficult for immigrant and racialized workers, particularly those in labour-intensive roles, and the COVID-19 pandemic exacerbated existing health inequities. Preparedness and response in high-risk industrial settings should therefore combine infection-control measures with multilingual communication, stronger worker protections, explicit anti-discrimination safeguards addressing ethnicity, language, and gender, and material supports that reduce the need to work while ill.

## Introduction

Workers in mass occupational settings such as meat processing plants were among the most negatively impacted by the COVID-19 pandemic.^1^ Mass occupational outbreaks occurred across the United States, Canada, Australia, Ireland, Germany, and the United Kingdom, driven by overcrowded production lines, high line speeds, cold working conditions, and loud environments that made physical distancing impractical and contributed to infections and fatalities, compromising workers’ health.^1,2^ Economic insecurity constrained workers’ ability to avoid these high-risk environments, while employer opacity about confirmed cases further amplified transmission risk within plants.^2^

Immigrant workers account for an estimated 52–70% of meat processing workforces in North America and Europe.^2^ In 2020, 45·4% of temporary foreign workers in Canada were employed in agriculture and agri-food sectors.^3^ Many meat processing plant workers face intersecting vulnerabilities, including language barriers, precarious immigration status, low wages, and limited healthcare access.^4^ These social determinants of health shape exposure, vulnerability, and health outcomes in high-risk occupational settings. These conditions reflect longstanding patterns of racialized labour stratification within large industrial workplaces that predate the pandemic.^5^

Meat processing plants in Alberta, Canada reported among the largest occupational COVID-19 outbreaks in North America, contributing significantly to community spread and raising concerns about workplace and community health.^2,6^ The structural conditions underlying these outbreaks, including occupational hierarchy, racialized labour concentration, income precarity, and limited worker agency, were documented and emphasized in the United Nations Special Rapporteur on Contemporary Forms of Slavery’s 2024 report, which described Canada’s Temporary Foreign Worker Program as institutionalising power asymmetries that prevent workers from exercising their rights.^7^ These structural vulnerabilities directly impacted workers’ ability to protect their health during outbreaks.

Existing studies of meat processing plants during COVID-19 have largely described outbreaks, epidemiological patterns and industrial transmission dynamics with limited attention to how health harms were distributed *within* workplaces along lines of occupational role, race, ethnicity, language, and income precarity.^1,6,8^ This gap limits the design of equitable occupational and public health interventions. We therefore conducted a community-engaged, exploratory sequential mixed-methods study guided by PROGRESS-Plus to examine how equity-relevant social and structural factors shaped self-reported COVID-19 diagnosis, physical and mental health, and experiences of discrimination among meat processing plant workers in Alberta, Canada.

## Methods

### Study Design, and Framework

This study used an exploratory sequential mixed-methods design, prioritising qualitative inquiry to identify equity-relevant mechanisms and then using a retrospective cross-sectional survey to quantify how those mechanisms were patterned across occupational and social strata.^9^ Grounded in community-based participatory research (CBPR), the design examined how social and structural determinants shaped workers’ lived experiences and perceived health during mass occupational outbreaks. The PROGRESS-Plus framework, encompassing place of residence, race identity, occupation, gender, religion, education, socioeconomic status, and social capital, plus additional context-specific factors, guided equity-relevant data collection and analysis throughout.^10^ Figure A illustrates the study phases. We use racialized identity to refer to self-reported racial categorization, recognizing race as a socially constructed process produced through structural and historical conditions. We reported findings in accordance with the Good Reporting of a Mixed Methods Study (GRAMMS) and Consolidated Criteria for Reporting Qualitative Research (COREQ) guidelines **(Supplements 1.1 and 1.2)**.^11,12^ The study was conducted in accordance with the Declaration of Helsinki and received ethics approval from the University of Calgary Research Ethics Board (REB20-1153).

### Participants, Plants, and Data Collection

We recruited participants between January and September 2021 from eleven meat processing plants across southern Alberta, Canada. Eligibility criteria were: (i) age ≥18 years; (ii) current or recent employment in a participating meat processing plant during the COVID-19 pandemic; and (iii) ability to provide informed consent in one of the study languages. Recruitment occurred through community outreach led by trained Community Scholars and partner organisations, using multilingual materials and multiple modes of participation (online, telephone-supported, and paper-based), described in detail elsewhere.^4^

For analytic purposes, we categorised plants by operating company and outbreak magnitude: two plants operated by Meat Processing Company 1 (MPC-1) experienced the largest regional outbreak and were grouped as MPC-1 (primary facility); all remaining plants were grouped as non-MPC-1.

We invited participants to complete one-on-one qualitative interviews (virtual or in-person) in their preferred language (English, Spanish, Filipino, Amharic, Tigrinya, Arabic, Oromo, or Dinka). Written consent was obtained in participants’ preferred language. Interviews were audio-recorded, transcribed verbatim, and translated into English by bilingual team members and Community Scholars, with spot-checking to preserve meaning in culturally specific expressions. Community Scholars then invited interview participants to complete an online or paper-based English survey **(Supplements 2.1 and 2.2)**. All interview participants also completed the survey; the qualitative sample was therefore fully nested within the quantitative sample. Participants could complete the survey independently or facilitated by Community Scholars in their preferred language, reducing linguistic and digital literacy barriers. We applied the Total Design Method to maximise response rates, following up with incomplete respondents at one, three, and seven-week intervals.^13^

The survey was adapted from validated instruments including the WHO COVID-19 Survey Tool,^14^ the Public Health Agency of Canada national case report form,^15^ and the Canadian Community Health Survey,^16^ with iterative pilot testing by Community Scholars to ensure cultural and linguistic relevance. It collected demographic characteristics (age, gender, racial identity, English proficiency, occupational role, immigration status), health-related outcomes, and equity-related measures including discrimination, income precarity, job insecurity, and perceived well-being **(Supplement 2.3)**.^17,18^

### Qualitative Analysis

We analysed qualitative data using inductive and deductive thematic analysis informed by the PROGRESS-Plus framework. Deductive codes captured how PROGRESS-Plus domains shaped perceived health inequities; inductive codes identified emergent themes related to biopsychosocial well-being. Each transcript was independently coded by two members of a six-person coding team, with different pairings used across transcripts to reduce individual bias; disagreements were brought to the full team for resolution by consensus, and the finalised codebook was applied across all transcripts. Thematic saturation was assessed iteratively, with coding continuing until no substantively new themes emerged. Community Scholars both coded and interpreted qualitative data, providing cultural and linguistic grounding for participant narratives. Their dual roles as community insiders and research team members positions their integration as a methodological strength that enhanced analytical credibility and informed our reflexive interpretive approach throughout. Qualitative data were managed and coded using NVivo (version 14; Lumivero, Denver, CO, USA).

### Quantitative Analysis

Descriptive statistics summarised demographic and occupational characteristics. Continuous variables were non-normally distributed and are reported as median and interquartile range (IQR). Differences in categorical variables across occupational groups were assessed using Pearson’s chi-square tests, while median scores were compared using the Kruskal-Wallis test. We estimated multivariable logistic regression models for three binary outcomes: self-reported COVID-19 diagnosis (yes/no), fair or poor self-rated general health (vs good/very good/excellent), and fair or poor self-rated mental health (vs good/very good/excellent). Everyday discrimination was measured using the Everyday Discrimination Scale (EDS), a validated nine-item instrument assessing the frequency of unfair treatment in daily life.^19^ A mean score was computed across all nine items (range 1–6, where 1=never and 6=almost every day), with higher scores indicating greater frequency of perceived discrimination. Items endorsed as ‘do not know or prefer not to answer’ were treated as missing and excluded from the mean calculation. Mean everyday discrimination score was modelled as a continuous outcome using linear regression.^17,19^

Occupational group, racialized identity, and facility (MPC-1 vs. non-MPC-1) were the primary exposures, selected a priori based on a conceptual model of workplace social stratification. For logistic regression models, adjusted odds ratios (aORs) with Wald 95% confidence intervals (CIs) are reported. For the linear regression model of everyday discrimination, unstandardized beta coefficients (β) with 95% CIs are reported, representing the mean difference in EDS score relative to the reference group. Reference categories were selected using qualitative findings to identify the most vulnerable groups: general labour, Black workers, and MPC-1. Models were estimated using complete-case analysis; “Prefer not to answer” responses were treated as missing. Variable-level missingness was assessed prior to model specification; predictors with missingness exceeding 15% were considered for exclusion from primary models. Prior to finalising the model specification, collinearity among predictors was assessed using a correlation matrix and generalised variance inflation factors. We reported model-specific sample sizes and variable-level missingness in accordance with STROBE guidance.^20^ Gender was collected as a self-reported variable, one participant who responded ‘I prefer not to answer’ was excluded from gender-stratified analyses. We conducted gender-stratified analyses in accordance with SAGER guidelines;^21^ however, gender was removed from the primary multivariable models due to collinearity with occupational group (primary exposure) and limited statistical power. All statistical analyses were conducted in R (version 4.3; R Core Team, R Foundation for Statistical Computing, Vienna, Austria). Forest plots and coefficient plots were generated using Python (version 3.11; Python Software Foundation, Wilmington, DE, USA).

### Integration

Integration occurred at the interpretation and reporting stages through joint displays, following established mixed-methods principles.^22,23^ We assessed convergence (qualitative and quantitative findings aligned), complementarity (one data type elaborated the other), and discordance (findings diverged). The full research team, including Community Scholars, used a whole-team consensus approach to interpret areas of discordance, including the divergence between qualitative accounts of workplace discrimination and quantitative EDS findings, recognising that generic instruments may not fully capture workplace-specific or retaliation-constrained forms of discrimination. Joint displays present qualitative and quantitative findings adjacently to facilitate transparent integration (Joint Displays 1-3).

## Results

### Qualitative Findings

Table 1 presents characteristics of the 36 interview participants. Most worked in general labour (25/36, 69·4%) and were predominantly Black (23/36, 63·9%) and male (28/36, 77·8%); language profiles aligned closely with occupational role, with Amharic and Tigrinya most common among general labourers and English most common among skilled labour and management. Half of participants held permanent resident status (18/36, 50·0%). Five themes emerged from the data **(Supplement 3.1)**: (1) unsafe work conditions; (2) discrimination; (3) lack of trust in safety measures; (4) COVID-19 impacts on family relationships; and (5) treatment and power hierarchy in large corporations.

**Table 1:** Interview Participant Characteristics.

| Characteristic | General Labour<br>n=25 (%) | Skilled Labour<br>n=5 (%) | Management n=4 (%) | Undeclared n=2 | Total n=36 |
| --- | --- | --- | --- | --- | --- |
| **Gender** |  |  |  |  |  |
| Men | 20 (80.0) | 3 (60.0) | 3 (75.0) | 2 (100.0) | 28 |
| Women | 5 (20.0) | 2 (40.0) | 1 (25.0) | 0 (0.0) | 8 |
| **Racialized Identity** |  |  |  |  |  |
| Black | 21 (84.0) | 0 (0.0) | 0 (0.0) | 2 (100.0) | 23 |
| White | 0 (0.0) | 0 (0.0) | 2 (50.0) | 0 (0.0) | 2 |
| Other | 3 (12.0) | 2 (40.0) | 1 (25.0) | 0 (0.0) | 6 |
| Undeclared | 1 (4.0) | 3 (60.0) | 1 (25.0) | 0 (0.0) | 5 |
| **Primary Language\*** |  |  |  |  |  |
| Amharic | 12 (48.0) | 0 (0.0) | 0 (0.0) | 2 (100.0) | 14 |
| Tigrinya | 8 (32.0) | 0 (0.0) | 0 (0.0) | 0 (0.0) | 8 |
| English | 1 (4.0) | 3 (60.0) | 3 (75.0) | 0 (0.0) | 7 |
| Tagalog/Filipino | 2 (8.0) | 2 (40.0) | 1 (25.0) | 0 (0.0) | 5 |
| Dinka | 1 (4.0) | 0 (0.0) | 0 (0.0) | 0 (0.0) | 1 |
| Spanish | 1 (4.0) | 0 (0.0) | 0 (0.0) | 0 (0.0) | 1 |
| **Immigration Status** |  |  |  |  |  |
| Permanent Resident | 17 (68.0) | 1 (20.0) | 0 (0.0) | 0 (0.0) | 18 |
| Citizen | 3 (12.0) | 2 (40.0) | 3 (75.0) | 1 (50.0) | 9 |
| Undeclared | 5 (20.0) | 2 (40.0) | 1 (25.0) | 1 (50.0) | 9 |
| *\*Though 8 languages were offered for interviews, no participants opted to be interviewed in Oromo or Arabic.* |  |  |  |  |  |

### Unsafe Work Conditions

Occupational role shaped health and well-being through exposure to overcrowding, inadequate safety protocols, and limited onsite information about confirmed infections. Participants described employers withholding information about active cases to sustain operations, leaving workers dependent on informal peer networks to assess their own risk. This pattern of deliberate opacity undermined workers’ ability to make informed decisions about their safety and that of their families.

> *“[W]e already felt like, it was like in the air, the fear, the uncertainty, because many people were already getting sick and the company didn’t give us any information. The company hid all that information, the company, they really deceived us for a long time because there were many positives and nobody knew. There were a lot of people who had already left work sick and no one knew.”* – Interview Participant 5

### Discrimination

Race, ethnicity, and language structured differential treatment within plant hierarchies. Participants described a workplace order in which country of origin and ethnicity determined role assignment and access to less physically demanding tasks, while language barriers constrained the ability to report unsafe conditions or advocate independently. Fear of employer retribution further suppressed disclosure, effectively silencing those most exposed to risk.

> *“Working at a meat processing plant, there is a lot of discrimination. I can only speak for MPC-1, because that is where I work… that is why they want minorities, because minorities are scared to speak up for themselves and for their rights… anybody who knows me at MPC-1, I speak English very well, and I am constantly speaking for myself or speaking up for my fellow co-workers, because they are too scared or they don’t know what to say.*“ – Survey Participant”

However, not all participants attributed differential treatment primarily to race. One participant challenged the framing of race as the central mechanism, instead pointing to immigration status and language as the more proximate drivers of vulnerability.

> *“I don’t believe colour plays a role at work… I never thought about my colour and my blackness as an issue… immigrant workers… we are living in Canada and we should take care of our business like Canadians and the work relationship has to change.”* Interview Participant 28

### Lack of Trust in Safety Measures

Even where safety protocols were visibly in place, participants expressed deep mistrust of their adequacy. Workers described being required to continue working while symptomatic, being denied requests for personal protective equipment, and observing that the physical demands and spatial constraints of production work made meaningful distancing structurally impossible.

> *“There were people asking for face masks and other personal protective equipment. And yet they’re saying that the company refused… there were people who, although they had COVID, they were called to come to work. Even though they had symptoms.”* – Interview Participant 14

### COVID-19 Impacts on Family Relationships

Infection and isolation produced major disruptions to family life. Participants described the emotional toll of separating from children and partners, and the damage to family relationships that resulted from prolonged quarantine and persistent fear of transmission within households after workplace exposures. These psychosocial harms extended well beyond the period of acute illness. Workers perceived this distancing as a source of isolation and sadness, as they found themselves unable to engage in the nurturing roles they typically held within their families.

> *“After screening, I was self-isolated from my children and my wife. That moment was totally against our traditional family care and nurturing tradition. That disease had terribly damaged our warm family relations.”* – Interview Participant 20

### Treatment and Power Hierarchy in Large Corporations

Hierarchal dynamics influenced participants’ experiences and treatment at work. This extended to the regulation of basic bodily functions, illustrating the structural conditions that constrained worker agency and dignity. Newer and lower-status workers described treatment that denied them the autonomy afforded to more senior colleagues, a dynamic that shaped psychological well-being and reinforced occupational stratification with direct health consequences.

> *“However, according to the company rule, if you want to use the washroom, the lead hand has to give you permission. Once you get the permission to use the washroom, they timed you in minutes and if you are late a bit, they take you to the office and make you sign, it is very stressful working condition.” –* Interview Participant 16

### Quantitative Findings

**Table 2** presents cohort characteristics by occupational group. Of 440 participants who registered, 191 completed the survey (43·4%); four were excluded for not reporting an occupational role, yielding a final analytic sample of 187: 123 general labourers (65·8%), 51 skilled labourers (27·2%), and 13 management staff (7%). The complete participant flow is presented in **Supplement 1.4**. Racialized identity and English proficiency differed markedly across occupational groups (both p<0·01), with Black workers comprising 55/123 (44·7%) of general labour compared with 16/51 (31·4%) of skilled labour and 1/13 (7·7%) of management, and excellent English proficiency reported by 29/123 (23·6%) of general labourers compared with 27/51 (52·9%) of skilled labour and 11/13 (84·6%) of management. Gender, income, and education level also varied significantly across occupational groups (all p<0·01); however, income precarity and facility did not differ meaningfully, nor did median age (38 years, IQR [32–44], 38 years IQR [32–50], 44 years IQR [36–50] for general labour, skilled labour and management respectively; Kruskal–Wallis p=0·44).

**Table 2:**
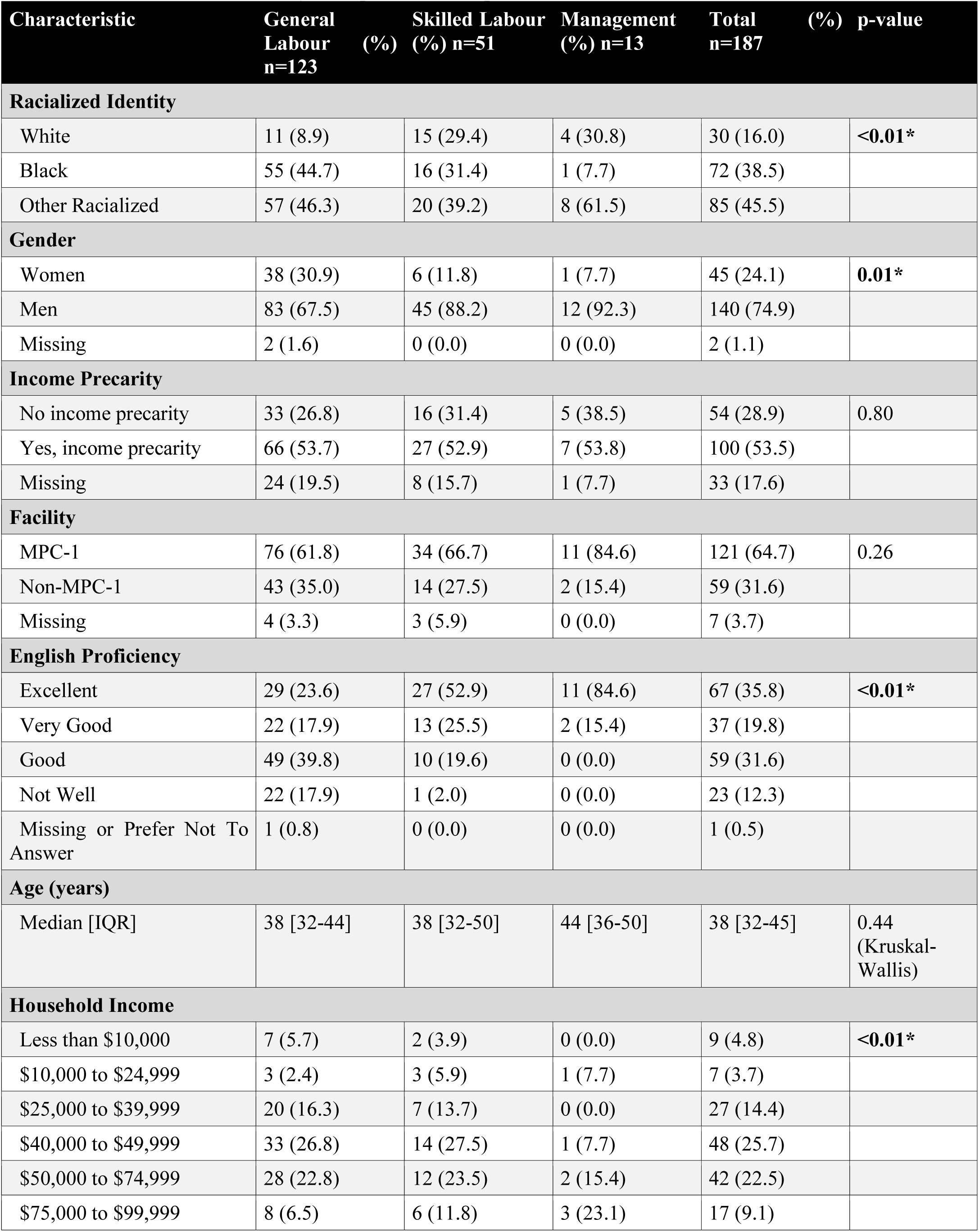
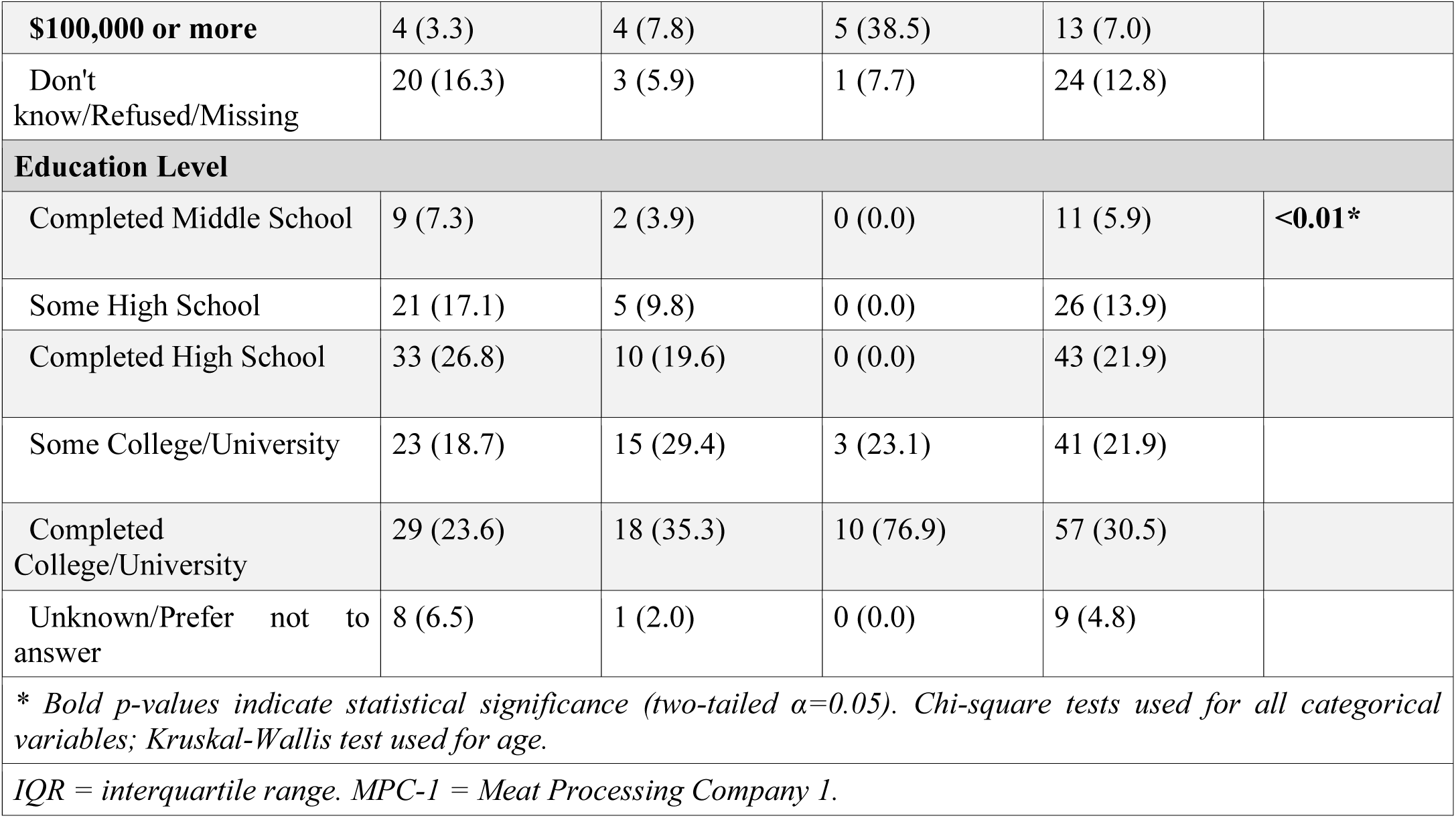
Cohort Characteristics by Occupational Group.

Variable-level missingness and model-specific sample sizes are reported in **Supplement 1.3** in accordance with STROBE guidance. Income precarity (money_diff_month) had the highest missingness of any model predictor (33/187, 17·6%) and was excluded from primary multivariable models on this basis; it is examined in a dedicated companion study.

Among 185 participants with complete COVID-19 outcome data, 55 (29·7%) reported a prior COVID-19 diagnosis. Unadjusted infection rates declined across the occupational hierarchy: 35·5% among general labourers, 21·6% among skilled labourers, and 7·7% among management (chi-square p=0·04, missing excluded). **Supplement 3.2** presents unadjusted outcomes across occupational groups. Most participants reported good or better self-rated general health (158/186, 84·9%) and mental health (139/182, 76·4%), with no significant unadjusted differences across occupational groups. Unadjusted everyday discrimination scores did not differ significantly across groups (Kruskal-Wallis median score p=0·49; **Supplement 3.3**).

In multivariable models (n=166–170; **Supplement 3.4**), skilled labour (aOR 0·38; 95% CI 0·15–0·89) and management (aOR 0·13; 95% CI 0·01–0·75) had lower odds of self-reported COVID-19 diagnosis compared with general labour; management estimates should be interpreted as directional given the small subgroup size (n=13). Workers at non-MPC-1 facilities also had significantly lower odds of COVID-19 diagnosis (aOR 0·29; 95% CI 0·12–0·67). Compared with Black workers, other racialized workers had substantially lower odds of fair or poor mental health (aOR 0·24; 95% CI 0·09–0·58) and fair or poor physical health (aOR 0·20; 95% CI 0·06–0·54). In linear regression models of mean everyday discrimination score, workers at non-MPC-1 facilities reported significantly lower mean discrimination scores compared with MPC-1 workers (β = –0·54; 95% CI –0·96 to –0·12; p=0·012). No significant associations were identified between occupational group or racialized identity and mean EDS score. Gender-stratified analyses among male participants largely replicated the primary model findings; no statistically significant associations were identified in the female stratum, consistent with limited statistical power given the small subsample size (n=38) and evidence of complete separation in several cells. Full gender-stratified results are presented in **Supplement 3.5**. Correlation analyses confirmed that binary facility classification adequately captured full facility variation (r=0·89) and identified moderate collinearity between gender and occupational group (r=0·21), supporting the use of binary facility categorisation and gender-stratified rather than adjusted analyses (**Supplement 3.6**). Adjusted model results for all outcomes are presented in Joint Displays 1-3.

## Integration

Integrated analysis revealed three overarching themes (Joint Displays 1-3). Across both data types, occupational role emerged as the primary structural determinant of COVID-19 exposure. Qualitative accounts of employer information-withholding, inadequate safety measures, and constrained worker agency were corroborated quantitatively: general labourers had significantly higher odds of self-reported COVID-19 diagnosis than those in skilled or management roles, suggesting that workplace hierarchy shaped not only lived experience but measurable health outcomes (**Joint Display 1**).

> *“[My employer] dropped the ball and … took forever for them to do it [shut down], they still kept running. They wouldn’t shut down, they were selfish and didn’t shut down and put all the stuff in place … because it’s freaking greedy … It’s brutal.” –* Interview Participant 11

Qualitative and quantitative findings on everyday discrimination showed partial discordance (**Joint Display 2**). Workers consistently described ethnicity, language, and seniority as structuring differential treatment within plants; however, no significant associations between occupational group or racialized identity and mean EDS score were identified in adjusted models, likely reflecting the limitations of a generic discrimination instrument in capturing workplace-specific, hierarchically-structured, and retaliation-constrained forms of unfair treatment where fear of reprisal shaped both experience and reporting. A point of convergence emerged at the facility level: workers at non-MPC-1 facilities reported significantly lower mean everyday discrimination scores than MPC-1 workers (β = –0·54; 95% CI –0·96 to –0·12), corroborating qualitative accounts of particularly pronounced differential treatment, power asymmetry, and employer opacity at MPC-1, and suggesting that discrimination frequency may vary meaningfully by facility. Notably, not all participants attributed differential treatment primarily to race; one participant pointed to immigration status and language as more proximate drivers of vulnerability, noting that immigrant workers were less likely to speak up and that English proficiency conferred a degree of protection. In this context, immigration status and language, which may coincide with racialized identity, were not collected or modelled separately, representing a limitation in the study’s capacity to disentangle the mechanisms through which discrimination operates in this setting.

> *“One thing it has to change is every employee has to be treated the same. There should not be discrimination based on color, race, ethnicity and gender, etc… The leaders at [MPC-1] treat people based on gender and ethnicity. There is a hierarchy based on country of origin and ethnicity.”* – Interview Participant 16

Qualitative and quantitative evidence converged on the psychological and social toll of working conditions, particularly for Black workers, who had significantly higher odds of fair or poor mental and physical health than other racialized workers. The lower odds of poor health among other racialized workers may partly reflect stronger within-group networks of mutual support and advocacy, where shared language, supervisory representation, and community solidarity provided a degree of protection not equally available to all groups. This pattern was supported by qualitative data, particularly from MPC-1, where differential access to in-group support was reported to structure both task assignment and day-to-day working conditions.

> *“ Our community is weaker. Our community does not help each other… I sense Indians and Philippines helping each other. I regret that I had not an opportunity to learn and communicate in English effectively.” –* Interview Participant 21
>
> *“At [MPC-1] if there is a Sudanese supervisor Sudanese are treated well, if you have a Philippines supervisor Filipinos are treated well and do easy jobs.” –* Interview Participant 16

Faith emerged as a coping mechanism providing comfort and a sense of agency amid uncertainty, particularly when institutional protections felt absent or unreliable.

> *“I trust GOD and would feel counseled.”* – Interview Participant 21
>
> *I didn’t feel safe, and we relied on God so the morning would come, and you are not sick. –* Interview Participant 42

One important area of discordance emerged around sex and gender: female workers described specific experiences of differential physical demands and unfair treatment not captured in quantitative group comparisons. This discordance likely reflects both the limitations of occupational group as the primary unit of stratification and the small female subsample (n=38), which precluded adequate statistical power to detect gender-specific associations. Discrimination was reported both based on gendered role expectations and biological sex, as one participant described targeted harassment and denial of basic accommodations following her return from maternity leave.

> *“I saw so many women crying because they could not express what is happening to them due to language barrier. It is very difficult to work there especially if you don’t have a lead hand or a supervisor from your country of origin or tribe.” –* Interview Participant *16*
>
> *“But when I returned from maternity leave… I felt the harassment, he wouldn’t even let me go to the bathroom, and if I went to the bathroom, he would let my work pile up there, he would do this only to me… I had to go to another human resources area, because I had already gone to human resources to report him many times, but no one listened to me.” –* Interview Participant 5

These findings point to the need for adequately powered, sex and gender-disaggregated analysis in future occupational health research in this setting **(Joint Display 3)**.

## Discussion

The COVID-19 pandemic did not create health inequity among immigrant and racialized meat processing plant workers in Alberta, instead it revealed and amplified existing structural inequities. Workers in general labour roles, who were disproportionately Black and of immigrant origin, experienced significantly higher odds of self-reported COVID-19 diagnosis, worse mental and physical health, and greater exposure to the occupational and social conditions that drove transmission. These gradients aligned precisely with the pre-existing occupational hierarchy, confirming that the pandemic intensified conditions it did not invent.

These findings extend a growing body of evidence that meat and poultry processing was a high-hazard environment before COVID-19, with crowded production lines, high line speeds, and cold and loud working conditions that made physical distancing structurally impractical.^2,6,24^ Meat processing plant workers have demonstrated substantially higher SARS-CoV-2 seroprevalence than workers in other food-system sectors, with infection risk specifically linked to these environmental characteristics and to working while symptomatic.^24^ Prior work has largely documented outbreak burden at the plant or population level; this study adds within-plant evidence showing that exposure and health consequences were distributed unequally across occupational strata, racialized identities, and income levels within the same workplaces. The marked outlier pattern associated with MPC-1, which experienced the region’s largest outbreak, suggests that plant-specific context and outbreak magnitude contributed to differential worker risk beyond the structural features common to all participating plants.

Racialized identity and language functioned as intersecting structural mechanisms rather than independent risk factors. Race and ethnicity shape health through differential access to resources, power, and protections, and in this study Black workers bore the greatest burden of poor mental and physical health independent of occupational role.^25,26^ Workers with lower English proficiency faced constrained access to timely health information and limited capacity to report unsafe conditions, particularly given the fear of employer retribution described throughout qualitative data. Economic insecurity drove presenteeism, compelling workers to continue working despite symptoms in the absence of adequate income protection.^27^ Gender emerged as an important factor of differential treatment qualitatively, with female workers describing disproportionate physical demands and targeted unfair treatment. Gender-stratified quantitative analyses were conducted but yielded no significant findings in the female stratum, most plausibly due to the small female subsample (n=38) and collinearity between gender and occupational group rather than absence of effect. Adequately powered, gender-disaggregated analyses in future research are needed to properly examine these associations.

These structural power asymmetries are not unique to this setting: the 2024 report of the UN Special Rapporteur on Contemporary Forms of Slavery described Canada’s Temporary Foreign Worker Program as “a breeding ground for contemporary forms of slavery, as it institutionalizes asymmetries of power that favour employers and prevent workers from exercising their rights.”^7^ Faith and social support networks also emerged as important sources of resilience among participants, consistent with evidence on religion and collective coping during COVID-19.^28,29^

These findings point to policy action at multiple levels. At the workplace level, employers in high-density essential industries should provide timely multilingual health and safety information directly to workers, ensure adequate ventilation and personal protective equipment, and involve workers meaningfully in occupational health decision-making rather than limiting communication to management channels. Employers should also implement enforceable anti-discrimination policies that explicitly address race, ethnicity, language, and occupational hierarchy, with independent oversight and clear protections for workers who report discriminatory treatment. At the regulatory level, occupational health and safety enforcement must include independent confidential reporting mechanisms that protect workers from retaliation, with meaningful accountability for non-compliance. At the labour policy level, paid sick leave and income protection are prerequisites for outbreak control: without them, income precarity compels presenteeism that sustains transmission and disproportionately harms those with the least power to refuse unsafe work. During this study, preliminary findings were used by the research team and community partners to successfully advocate for paid sick leave at one large facility where protections had previously been absent, and later to successfully advocate for vaccine prioritization of all meat processing plant and other high-risk occupational settings across Alberta.^4^ This demonstrates that community-engaged research can directly drive workplace policy change during a health emergency.^30^ These protections should be treated as baseline requirements in high-risk industrial settings and extended to all workers regardless of immigration or employment status.^4^

## Limitations

This study has several limitations. Survey completion was 43·4%, raising the possibility of selection bias if participation differed by occupational role, language, or prior experience of harm; those most exposed or most fearful of reprisal may have been least likely to participate, potentially underestimating the magnitude of inequities reported here. Outcomes were self-reported and cross-sectional, limiting causal inference and introducing the possibility of recall and social desirability bias. Although community scholar-led data collection, participant language matching, and company anonymization were used to mitigate the influence of non-disclosure agreements and fear of employer retribution, residual response suppression cannot be excluded. Estimates for the management subgroup (n=13) were imprecise and should be interpreted as indicating direction and magnitude rather than precise effects. Translation across eight languages, while carefully managed with spot-checking by Community Scholars, may have introduced some loss of nuance. The Everyday Discrimination Scale may not fully capture workplace-specific, hierarchy-embedded, or retaliation-constrained forms of discrimination, which likely accounts for the discordance between qualitative and quantitative findings on this outcome. Further, statistical significance should be interpreted in the context of effect size, prior hypotheses, and qualitative triangulation rather than as confirmatory hypothesis testing. Gender-stratified analyses were conducted in accordance with SAGER guidelines but yielded no significant findings, most likely reflecting limited statistical power, complete separation in several cells, and collinearity between gender and occupational group rather than a true absence of effect. The gendered experiences described qualitatively warrant dedicated investigation in future studies with adequate female representation.

## Conclusion

In Alberta meat processing plants, COVID-19 harms were patterned by occupational hierarchy and racialization, reflecting the amplification of structural inequities that predated the pandemic. Workers in frontline labour roles, disproportionately Black workers, bore the greatest burden of infection, psychological harm, and social disruption while having the least power to protect themselves. Equitable pandemic preparedness in essential industrial workplaces requires adequate ventilation and personal protective equipment, paid sick leave and income protection, and multilingual worker communication. It also demands independent reporting mechanisms free from retaliation risk with enforceable accountability measures and genuine worker participation in occupational health decision-making. Finally, dismantling racialized occupational stratification requires enforceable equity standards in occupational health and labour legislation, mandatory disaggregation of workplace health and safety data by racialized identity and occupational role, and structural accountability for employers who concentrate racialized workers in the highest-risk positions. These are not exceptional crisis measures. They are baseline conditions for workplaces that depend on racialized labour.

## Declarations

### Ethics approval and consent to participate

This study was conducted in accordance with the Declaration of Helsinki and received ethics approval from the University of Calgary research ethics board (REB20-1153).

### Consent for publication

Not applicable.

### Data sharing statement

De-identified quantitative data are available upon reasonable request. Qualitative transcripts are not available for sharing due to participant confidentiality and ethics constraints.

### Authors’ contributions

**Gabriel E. Fabreau**: Conceptualization, Methodology, Supervision, Project administration, Validation, Writing – original draft, Writing – review & editing. **Kevin Pottie:** Conceptualization, Methodology, Supervision, Project administration, Validation, Writing – original draft, Writing – review & editing. **Mohammad Yasir Essar:** Data curation, Formal analysis, Investigation, Writing – original draft, Writing – review & editing. **Eric Norrie:** Data curation, Formal analysis, Investigation, Writing – original draft, Writing – review & editing. **Edna Ramirez Cerino:** Data curation, Formal analysis, Investigation, Writing – original draft, Writing – review & editing. **Minnella Antonio:** Data curation, Formal analysis, Investigation, Writing – original draft, Writing – review & editing. **Ammar Saad:** Data curation, Formal analysis, Investigation, Writing – original draft, Writing – review & editing. **Mussie Yemane:** Data curation, Formal analysis, Investigation. **Linda Holdbrook:** Data curation, Formal analysis, Investigation. **Adanech Sahilie:** Data curation, Formal analysis, Investigation. **Michael Youssef:** Data curation, Formal analysis, Investigation. **Nour Hassan:** Data curation, Formal analysis, Investigation. **Olivia Magwood:** Writing – original draft, Writing – review & editing. **Samuel T. Edwards:** Writing – original draft, Writing – review & editing. **Denise Spitzer:** Writing – review & editing. **Annalee Coakley:** Writing – original draft, Writing – review & editing. All authors approved the final manuscript.

## Declaration of generative AI and AI-assisted technologies in the writing process

During the preparation of this work the author(s) used artificial intelligence (ChatGPT, OpenAI, San Francisco, CA, USA) to improve grammar and readability. After using this tool/service, the author(s) reviewed and edited the content as needed and take(s) full responsibility for the content of the published article.

## Supporting information

COREQ Checklist

## Acknowledgements

We would like to acknowledge the immigrant and racialized workers who participated in our study. We value your time and trust, and hope that this work can inform meaningful change.

---

**Figure A.**
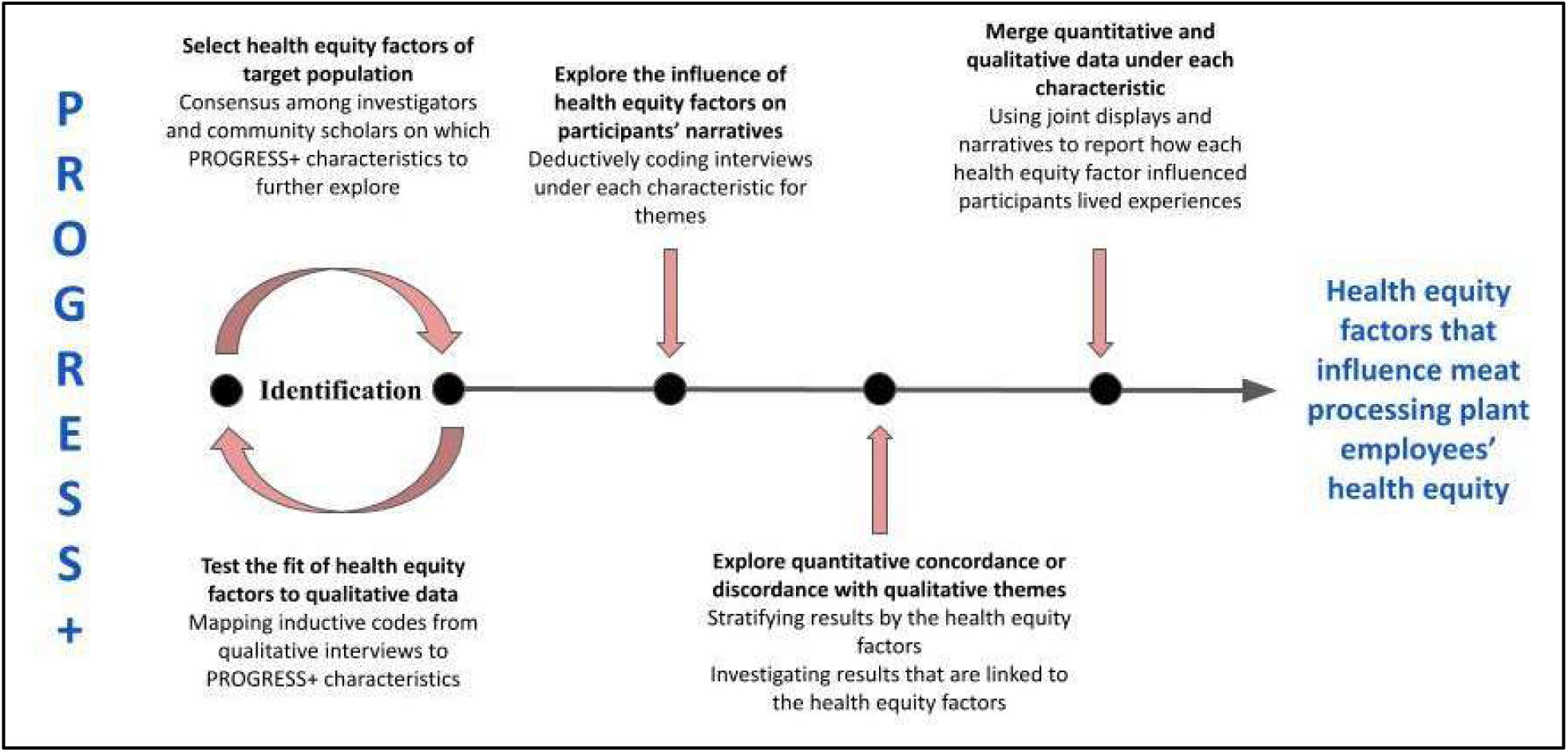
Conceptual Framework and Mixed Methods analysis plan

**Fig. Joint Display 1.**
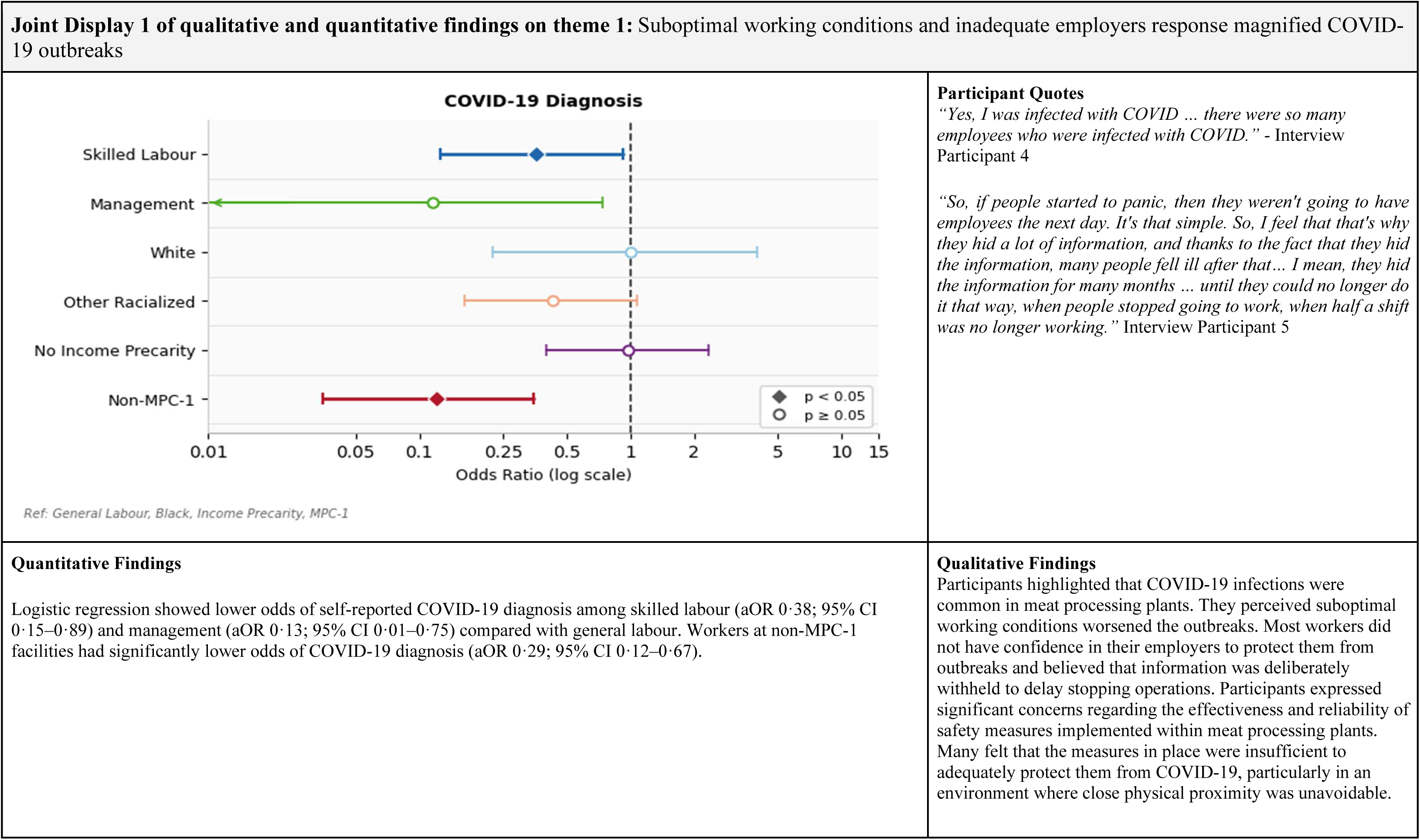
Adjusted odds ratios (aORs) with 95% confidence intervals from multivariable logistic regression models assessing associations between occupational group, racialized identity, and facility (MPC-1 vs. non-MPC-1) and self-reported COVID-19 diagnosis. Odds ratios are displayed on a logarithmic scale. The vertical line indicates the null value (OR=1). Reference groups were General Labour (occupational group), Black (racialized identity), and MPC-1 (facility). Confidence intervals were estimated using the Wald method. Apparent asymmetry of confidence intervals around point estimates on the log scale reflects numerical instability of the Wald approximation in small subgroups and should be interpreted with caution.

**Fig. Joint Display 2.**
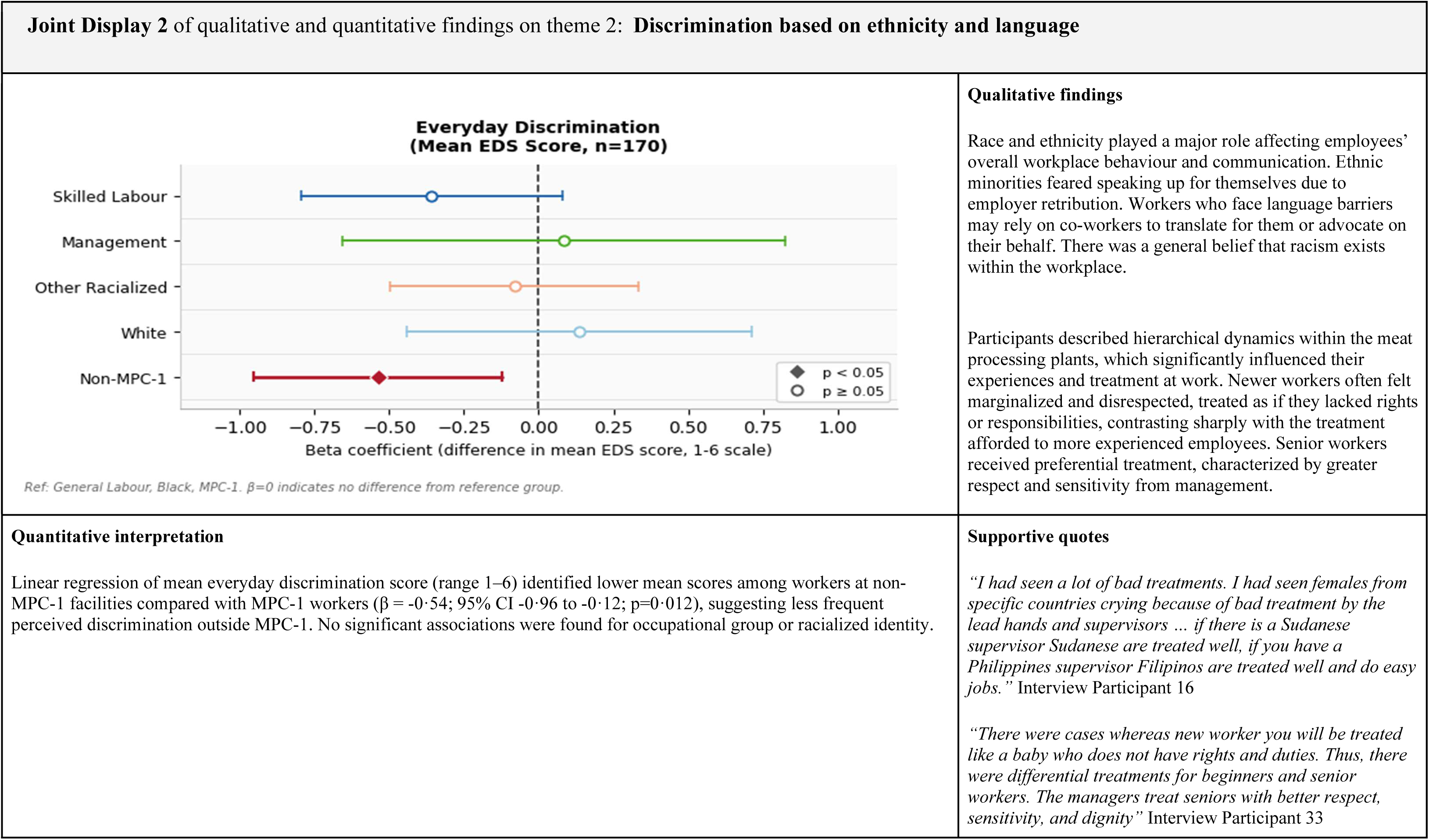
Unstandardised beta coefficients (β) with 95% confidence intervals from a linear regression model assessing associations between occupational group, racialized identity, and facility (MPC-1 vs. non-MPC-1) and mean everyday discrimination score (range 1–6, higher = more frequent discrimination). The vertical line indicates the null value (β=0). Reference groups were General Labour (occupational group), Black (racialized identity), and MPC-1 (facility). Confidence intervals are 95% CIs.

**Fig. Joint Display 3.**
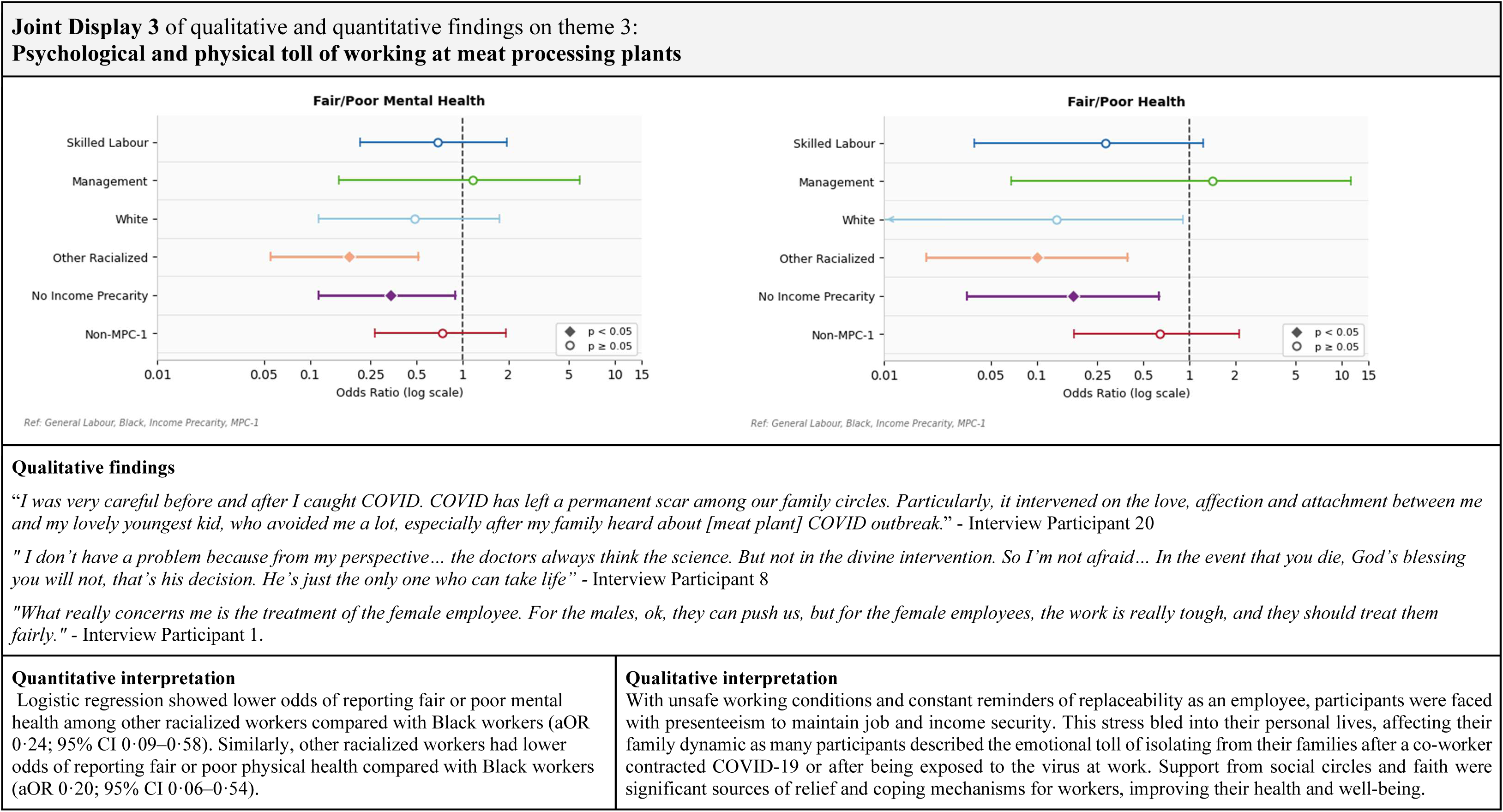
Adjusted odds ratios (aORs) with 95% confidence intervals from multivariable logistic regression models assessing associations between occupational group, racialized identity, and facility (MPC-1 vs. non-MPC-1) and outcomes of fair or poor mental health and fair or poor physical health. Odds ratios are displayed on a logarithmic scale. The vertical line indicates the null value (OR=1). Reference groups were General Labour (occupational group), Black (racialized identity), and MPC-1 (facility). Confidence intervals were estimated using the Wald method. Apparent asymmetry of confidence intervals around point estimates on the log scale reflects numerical instability of the Wald approximation in small subgroups and should be interpreted with caution.

