## Supplement 1 for "Occupational hierarchy, racialization, and COVID-19 health outcomes among meat processing plant workers in Alberta: a community-engaged mixed-methods study"

**Supplement 1: Reporting Guideline Checklists**

**Supplement 1.1: GRAMMS — Good Reporting of a Mixed Methods Study**

| Good Reporting of A Mixed Methods Study (GRAMMS) |  |
| --- | --- |
| Reporting Item | Where in Manuscript |
| (1) Describe the justification for using a mixed methods approach to the research question | Introduction: Page 6 |
| (2) Describe the design in terms of the purpose, priority and sequence of methods | Methods: Page 6-7 |
| (3) Describe each method in terms of sampling, data collection, and analysis. | Methods: Page 7-8 |
| (4) Describe where integration has occurred, how it has occurred, and who has participated in it. | Methods and Results: Page 9 |
| (5) Describe any limitation of one method associated with the present of the other method. | Discussion: Page 18 |
| (6) Describe any insights gained from mixing or integrating methods. | Discussion: Page 18-19 |

**Supplement 1.2: COREQ — Consolidated Criteria for Reporting Qualitative Research**

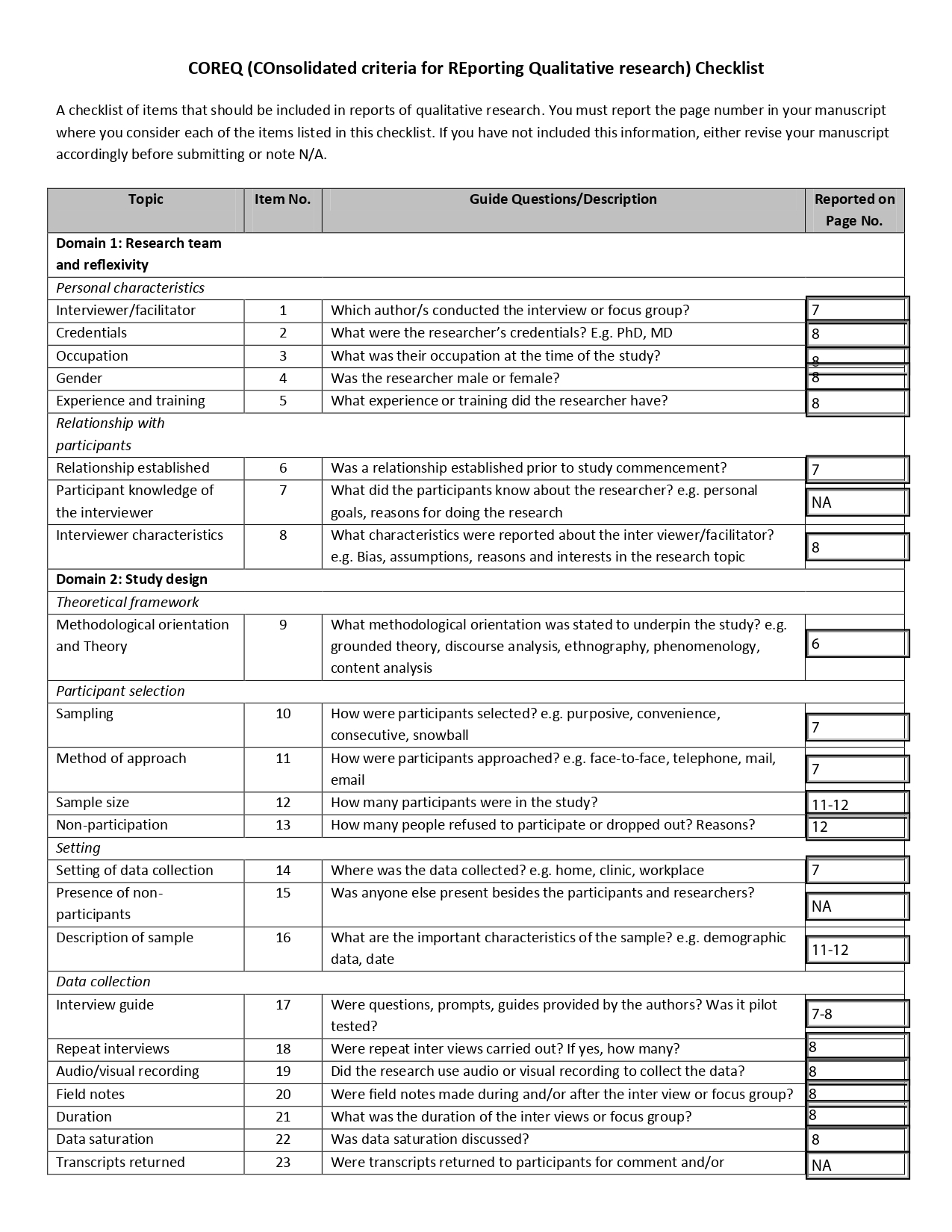

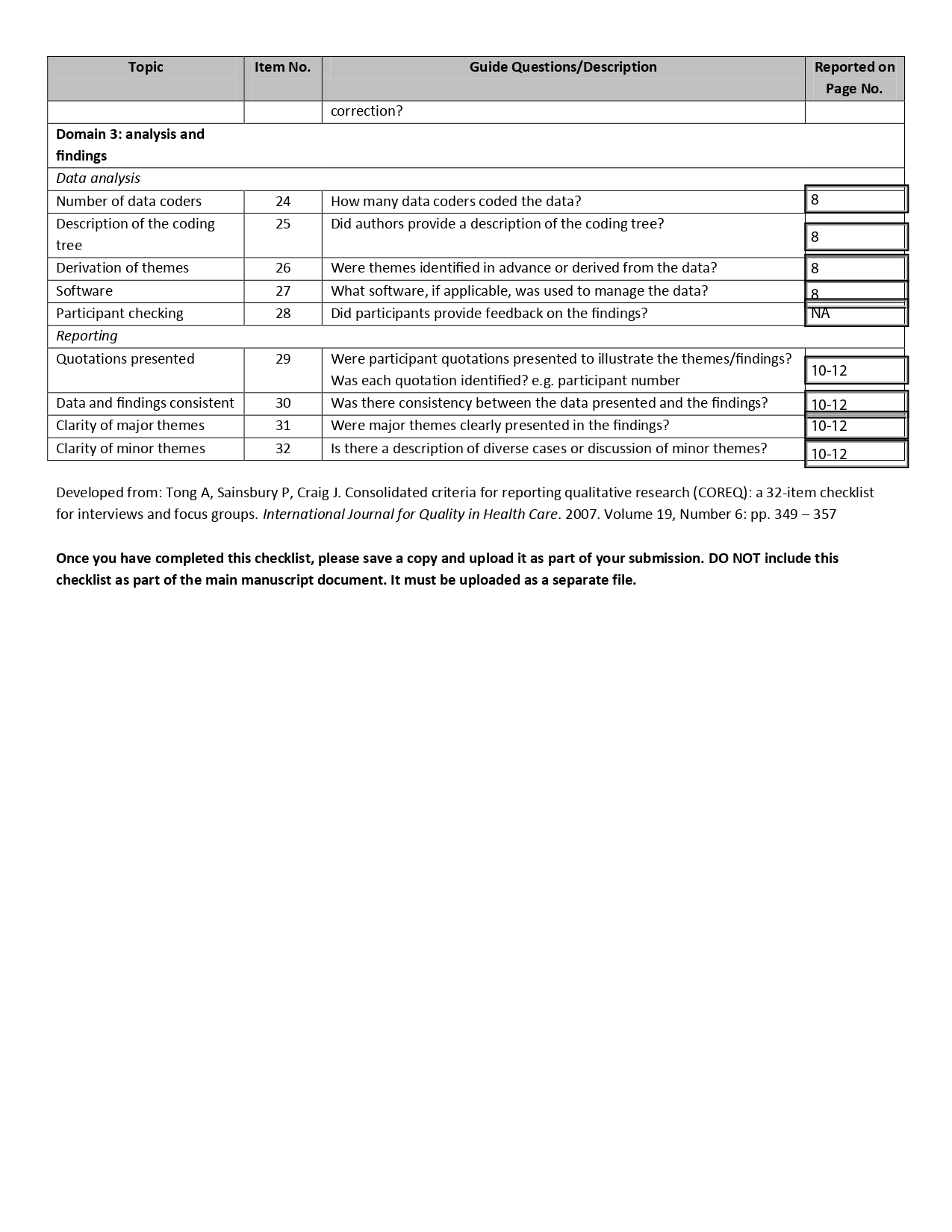

**Supplement 1.3: STROBE — Sample Size Attrition and Variable-Level Missingness**

*Sample-size flow and missingness reporting in line with STROBE recommendations.*

| **Part A: Study Attrition — From Registered Participants to Analytic Sample** | | | |
| --- | --- | --- | --- |
| **Filter Step** | **Reason** | **n Remaining** | **n Lost** |
| Raw dataset | All registered participants | 191 | — |
| Remove invalid occupational group | occ_group = '0' or missing | 187 | 4 |
| Remove missing racialized identity | racialized = NA | 177 | 10 |
| Remove missing facility | Facility (MPC-1 vs. non-MPC-1) = NA | 170 | 7 |
| **Base analytic sample** | **All primary predictors complete** | **170** | **—** |
| **Remove missing income precarity*** | **money_diff_month = NA or 'Prefer not to answer'** | **140** | **30** |
| **Sensitivity analysis sample** | **Income precarity included (sensitivity analysis only)** | **140** | **—** |
| **Income Precarity excluded from primary models due to 17.6% missingness; a separate paper examines income precarity in this cohort.* | | | |

| **Part B: Model-Specific Analytic Sample Sizes (Primary Models, base n=170)** | | | |
| --- | --- | --- | --- |
| **Outcome** | **Analytic n** | **Lost from Base** | **Reason for Loss** |
| Everyday Discrimination | 170 | 0 | No outcome missingness |
| Fair/Poor Health | 169 | 1 | 1 'Prefer not to answer' on health scale |
| COVID-19 Diagnosis | 168 | 2 | 2 missing COVID-19 diagnosis responses |
| Fair/Poor Mental Health | 166 | 4 | 4 'Prefer not to answer' or 'Do not know' on meant health |

| **Part C: Variable-Level Missingness (from base n=187, post occupational group filter)** | | | | | | |
| --- | --- | --- | --- | --- | --- | --- |
| **Variable** | **Role** | **Missing (NA)** | **Prefer Not to Answer / DK** | **Total Lost** | **n Remaining** | **% Missing** |
| Age (estimated_age) | Demographic | 88 | 0 | **88** | 99 | **47.1%** |
| Racialized identity (racialized) | Predictor | 10 | 0 | 10 | 177 | 5.3% |
| Facility (MPC-1 vs. non-MPC-1) (binary facilities) | Predictor | 7 | 0 | 7 | 180 | 3.7% |
| Income Precarity (money_diff_month) | Predictor* | 18 | 15 | **33** | 154 | **17.6%** |
| Sex (sex) | SAGER variable | 1 | 1 | 2 | 185 | 1.1% |
| COVID-19 Diagnosis (covid) | Outcome | 2 | 0 | 2 | 185 | 1.1% |
| Self-rated Physical Health (health scale) | Outcome | 0 | 1 | 1 | 186 | 0.5% |
| Self-rated Mental Health (meant health) | Outcome | 0 | 5 | 5 | 182 | 2.7% |
| Everyday Discrimination Scale (ed1-ed9) | Outcome (EDS) | 0 | 0 | 0 | 187 | 0.0% |
| *Dark grey: ≥10 participants lost (≥5.3% missingness). *Income Precarity excluded from primary models. DK = Do not know.* | | | | | | |
| *SAGER sex-stratified analyses: Male n=130, Female n=38 (after sex missingness exclusions from base n=170).* | | | | | | |
| *l models use complete-case analysis. 'Prefer not to answer' responses treated as missing throughout.* | | | | | | |

**Supplement 1.4: Participant Flow Diagram**

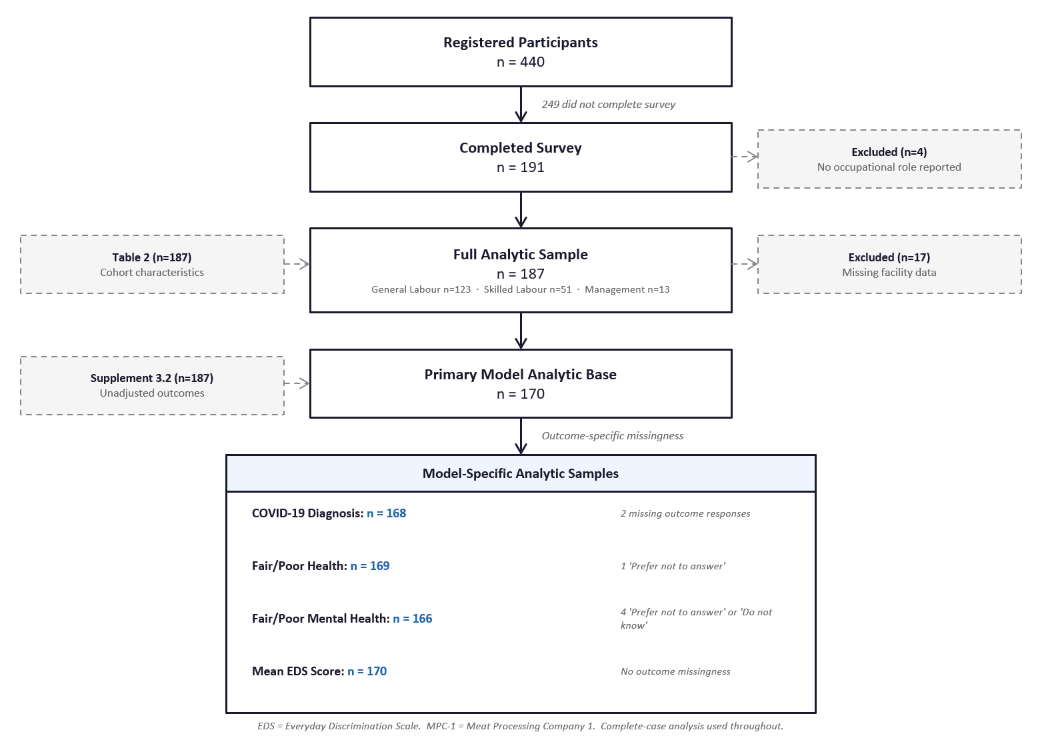
