## Supplement 2 for "Occupational hierarchy, racialization, and COVID-19 health outcomes among meat processing plant workers in Alberta: a community-engaged mixed-methods study"

**Supplement 2: Survey and Interview Guides**

**Supplement 2.1: Survey Instrument**

*The survey instrument administered to participants. Both facilitated and self-administered versions are included.*

*** Introduction ***

For facilitated surveys: Thank you again for agreeing to do this survey today. Just to re-iterate, you may choose to skip any questions you do not want to answer, just let me know and we will move along. If you need to take a break for water or washroom in the middle of the survey, please let me know. If you do not have any further questions, I would like to begin by collecting some basic information.

For non-facilitated surveys: Thank you again for agreeing to do this survey. Just to re-iterate, you may choose to skip any questions you do not want to answer. This survey will take approximately 30 minutes but will be active for 14 days. We will begin by collecting demographic information

**Date of Birth: __ __/__ __/ __ __ __ __**

DD	MM	YEAR

o Unknown

**Estimated age (if DOB is unknown):** _______________

**Postal Code:** ____________________________

**Record ID** (for RedCap)**: ___**

**Date of Collection: __ __/__ __/ __ __ __ __**

DD	MM	YEAR

**Interviewer initials: ______________**

**This survey is being completed by (Select ONE):**

o Meat processing plant employee	o Community member

o Other food industry employee (i.e., UCFW member) o Other ___________________

**Notes:**

[DEMOGRAPHICS]

These next set of questions ask about your sex, gender, ethnic background, children, languages you speak, and your past employments and current employment.

The following questions are about sex at birth and gender. Sex refers to sex assigned at birth. Gender refers to current gender which may be different from sex assigned at birth and may be different from what is indicated on legal documents.

SEX AND GENDER (GDR)

1. **What was your sex at birth?**

1: Male

2: Female

8: I prefer not to answer

9: Don’t know

2. **What is your gender?**

1: Man

2: Woman

3: Nonbinary

4: Prefer not to disclose

5: Other _____________

8: I prefer not to answer

9: DK

3. **What is your postal code?**

____________

4. **You may belong to one or more racial or cultural groups on the following list. Do you identify with? Please check all relevant identities**

01: White

02: South Asian (e.g., East Indian, Pakistani, Sri Lankan)

03: Chinese

04: Black Caribbean (e.g. Barbadian, Jamaican)

05: Black African (e.g. Ghanian, Somali, Kenyan, Sudanese, Egyptian)

06: Filipino

07: Latin American

08: Arab

09: Southeast Asian (e.g., Vietnamese, Cambodian, Malaysian, Laotian)

10: West Asian (e.g., Iranian, Afghan)

11: Korean

12: Japanese

13: First Nations

14: Metis

15: Inuit

16: Other – Specify___________________________

98: I prefer not to answer

99: DK

5. **What language do you speak most often at home?** ______________________
6. **How well would you say you speak and understand English? [English Proficiency].**

o Excellent

o Very good

o Good

o Not well

o Not well at all

o Prefer not to answer

7. **What is the highest level of education you have completed?**

o Completed Middle School

o Some High School

o Completed High School

o Some College/ University

o Completed College/ University

o Unknown

o Prefer not to answer

[FAMILY QUESTIONS]

The next questions are about your family and household. Just a reminder that your responses will be kept COMPLETELY CONFIDENTIAL and will not be shared. Your honesty in answering these questions will assist us in obtaining a clearer picture of the employees at your workplace.

8. **What is your current marital status?**

o Single (Never legally married)

o Widowed

o Divorced

o Remarried

o Legally married (and not separated)

o Separated, but still legally married

o Common-law partner (live together as a couple but are not legally married)

o Unknown

o Prefer not to answer

9. **Who lives in your household besides yourself?** *Choose as many as apply*

o l live alone

o I live with others

o l live with children under 18 years

o I live with people over 65 years

o l live with people who have chronic disease (s)

o None of the above

10. **How many individuals are with you in your household: ______** people
11. **How many individuals in your household are 65 years of age or older:**
12. **How many children under 18 years currently live in your household? _________**
    a. What are their ages? _________

YOUR HEALTH

[GENERAL HEALTH]

The next questions are about your health. By health, we mean not only the absence of disease or injury but also physical, mental and social well-being.

13. **In general, would you say your health is...?**

1: Excellent

2: Very good

3: Good

4: Fair

5: Poor

8: I prefer not to answer

9: DK

14. **Using a scale of 0 to 10, where 0 means "Very dissatisfied" and 10 means "Very satisfied", how do you feel about your life as a whole right now?**

**Very dissatisfied	Very satisfied	DK**

**0	1	2	3	4	5	6	7	8	9	10	88**

15. **In general, would you say your mental health is...?**

1: Excellent

2: Very good

3: Good

4: Fair

5: Poor

8: I prefer not to answer

9: DK

16. **Thinking about the amount of stress in your life, would you say that most of your days are...?**

1: Not at all stressful

2: Not very stressful

3: A bit stressful

4: Quite a bit stressful

5: Extremely stressful

8: I prefer not to answer

9: DK

**The next questions are a comparison of your workdays before and during COVID-19 pandemic**

17. **Before the COVID-19 pandemic, would you say that most days at work were...?**

1: Not at all stressful

2: Not very stressful

3: A bit stressful

4: Quite a bit stressful

5: Extremely stressful

8: I prefer not to answer

9: DK

18. **Since COVID-19 emerged and presently, would you say that most days at work were...?**

1: Not at all stressful

2: Not very stressful

3: A bit stressful

4: Quite a bit stressful

5: Extremely stressful

8: I prefer not to answer

9: DK

19. **How would you describe your sense of belonging to your workplace? Would you say it is...?**

1: Very strong

2: Somewhat strong

3: Somewhat weak

4: Very weak

8: I prefer not to answer

9: DK

20. **How would you describe your sense of belonging to your local community? Would you say it is...?**

1: Very strong

2: Somewhat strong

3: Somewhat weak

4: Very weak

8: I prefer not to answer

9: DK

IMMIGRATION QUESTIONS

If you have immigrated to Canada, we would like to ask you questions about your immigration status, your home country, and your arrival in Canada. Just a reminder that your responses will be kept COMPLETELY CONFIDENTIAL and will not be shared.

**Acronyms**:

GAR – Government-Assisted Refugee; CR1 – convention refugees, category 1

PSR – Privately-Sponsored Refugee; CR3 – convention refugees, category 3

BVOR – Blended Visa Office-Referred	TFW – Temporary Foreign worker

21. **Citizenship / Immigration status:**

o Canadian citizen

o Landed immigrant/ Permanent Resident

o Temporary Foreign Worker

o Government Assisted Refugee – GAR/CR1

o Privately Sponsored Refugee – PSR/CR3

o Refugee Claimant

o BVOR

o Permanent resident

o Unknown

o Other: ____________________________

o Prefer not to answer

22. **Please specify refugee claimant status:**

O Applied

O Accepted

O Rejected or appealed

O Awaiting Deportation

23. **If you have immigrated to Canada, what is your country of origin?** ____________________

o Prefer not to answer

24. **What date did you arrive in Canada? Please leave blank if you do not know or prefer not to answer	__ __/__ __/ __ __ __ __**

DD	MM	YEAR

Work status – We would like to ask you about your work or job.

25. **What is your current employment status at the meat processing plant?**

o Full-time employee (working 40 or more hours/week)

o Part-time employee

o Casual employee

o Temporary employee

o Fixed term/contract employee

o Training/Apprentice

o Other: ____________________

26. **What is your current position at the meat processing plant?**

o General Production

o Fabrication Floor

o Shipping

o Skilled Production

o Maintenance

o Meat cutter

o Manager/Supervisor

o Other ___________________

27. **How long have you worked at your current place of work?** ____ years _____ months
28. **How many hours on average do you work per week?**

o Less than 10 hours/ week

o 10 – 20 hours/week

o 20 – 30 hours/week

o 30 – 40 hours/week (full-time)

o More than 40 hours/week

29. **If you missed time at work due to sickness or were required to isolate at home, did you receive or have access to sick pay?**

o Yes

o No

o Don’t know/Unsure

30. **Were you offered or did you receive bonus pay for working during the COVID-19 outbreak at your job?**

o Yes, offered but did not receive bonus pay

o Yes, received bonus pay

o No

o Don’t know/Unsure

CHRONIC CONDITIONS

We would like to know about "long-term health conditions" which are expected to last or have already lasted 6 months or more and that have been diagnosed by a health professional.

31. **Has your doctor or health professional told you that you have any of the following in the last 12 months?:**

o Asthma

o High blood pressure or Hypertension

o Heart disease

o Diabetes

o Chronic bronchitis, Emphysema, or COPD

o Obesity or excessive weight

o High blood cholesterol or lipids

o Effects of a stroke

o Cancer

o Kidney disease

o Arthritis, (for example osteoarthritis, gout or any other type)

o Fibromyalgia

o Chronic fatigue

o Mood disorder such as depression, bipolar disorder, mania or dysthymia?

o Anxiety disorders such as phobia, Obsessive-compulsive disorder or a panic disorder?

O No chronic health conditions

o Other ______________________

**SMOKING –** The next questions are about cigarette smoking:

32. At the present time do you smoke cigarettes every day, occasionally, or not at all?

O Everyday

O Occasionally

O Not at all

O Prefer not to answer

33. In the past 30 days, did you smoke any cigarettes?
    o Yes

O No

O Prefer not to answer

34. If you were a regular smoker, when did you stop smoking? Was it…
    o Less than 1 year ago

O 1 year to less than 2 years ago

O 2 years to less than 3 years ago

O 3 or more years ago

O I did not stop smoking

O I have never smoked

O Don’t know

O Prefer not to answer

COVID-19 PERSONAL EXPERIENCE

We would like to ask you specifically about your experience with and understanding of COVID-19.

35. **To your knowledge, are you, or have you been, infected with COVID-19?**

O No

O Yes

If yes, was it:

O Mild? – did not need to go to a hospital

O Severe? – needed to go to the emergency department or was hospitalized

O Asymptomatic or no symptoms?

O Pre-symptomatic? (recently infected but don’t have any symptoms yet)

Was the diagnosis confirmed by a test:

O Confirmed by a test?

O Not confirmed by a test?

36. **When were you diagnosed with COVID-19? (month/year) _________**
37. **What symptoms did you develop? Check all that apply**
    - Cough
    - Shortness of breath
    - Temperature equal or over 38 C
    - Fever
    - Chills
    - Fatigue
    - Muscle or body aches
    - New loss of smell or taste
    - Headache
    - Gastrointestinal symptoms
    - Feeling very unwell
    - Other? ____________
38. **How do you think you were infected with COVID-19?**
    a. Work
    b. Home
    c. Community
    d. Unsure/ DK
    e. Other ________________
39. **Was anyone else infected in your household ? (Yes/No)**
    a. If yes, Who? ______________ How Many? __________ people
    b. If no, Why/how was this prevented? _________________
40. **Do you know anyone who was admitted to the hospital or intensive care unit (ICU) due to COVID-19?**

O No

O Yes

o Don’t know/Unsure

**If yes,** who were they? ____________________________

41. **Do you know anyone who has died from COVID-19?**

O No

O Yes

o Don’t know/Unsure

**If yes,** who were they? ____________________________

HEALTH LITERACY
We would like to ask you about your thoughts about COVID-19 and where you get CODI-19 information.

**How easy or difficult would you say it is to…?:**

42. **…find the information you need related to COVID-19?**

**Very difficult	Very easy	DK**

**0	1	2	3	4	5	6	7	8	9	10	88**

43. **…understand information about what to do if you think you have COVID-19?**

**Very difficult	Very easy	DK**

**0	1	2	3	4	5	6	7	8	9	10	88**

44. **…judge if the information about COVID-19 in the media is reliable?**

**Very difficult	Very easy	DK**

**0	1	2	3	4	5	6	7	8	9	10	88**

45. **…understand restrictions and recommendations of health authorities regarding COVID-19?**

**Very difficult	Very easy	DK**

**0	1	2	3	4	5	6	7	8	9	10	88**

46. **…follow the recommendations on how to protect yourself from COVID-19?**

**Very difficult	Very easy	DK**

**0	1	2	3	4	5	6	7	8	9	10	88**

47. **…understand recommendations about when to stay at home from work/school, and when not to?**

**Very difficult	Very easy	DK**

**0	1	2	3	4	5	6	7	8	9	10	88**

48. **…understand recommendations about when to engage in social activities, and when not to?**

**Very difficult	Very easy	DK**

**0	1	2	3	4	5	6	7	8	9	10	88**

PREPAREDNESS & PERCEIVED SELF-EFFICACY

**Next, we would like to know about you own practices related to COVID-19.**

49. **I know how to protect myself from COVID-19:**

**Very much so	Not at all	DK**

**0	1	2	3	4	5	6	7	8	9	10	88**

50. **Avoiding an infection with COVID-19 in the current situation is easy…**

**Not at all	Very much so	DK**

**0	1	2	3	4	5	6	7	8	9	10	88**

PREVENTION – OWN BEHAVIORS

51. **During the last 7 days, which of the following measures have you taken to prevent infection from COVID-19?**

*Choose as many as apply*

| o Frequently washed my hands with soap and water for at least 20 seconds | Not at all - - - - - - - - - - - - - Very much so - - - DK/Refused |
| --- | --- |
| o Avoided touching my eyes, nose, and mouth with unwashed hands | Not at all - - - - - - - - - - - - - Very much so - - - DK/Refused |
| o Used hand sanitizer to clean hands when soap and water were not available | Not at all - - - - - - - - - - - - - Very much so - - - DK/Refused |
| o Avoided a social event I wanted to attend | Not at all - - - - - - - - - - - - - Very much so - - - DK/Refused |
| o Stayed at home from work/school | Not at all - - - - - - - - - - - - - Very much so - - - DK/Refused |
| o Used antibiotics to prevent or treat COVID-19 | Not at all - - - - - - - - - - - - - Very much so - - - DK/Refused |
| o Wore a mask in public | Not at all - - - - - - - - - - - - - Very much so - - - DK/Refused |
| o Ensured physical distancing in public | Not at all - - - - - - - - - - - - - Very much so - - - DK/Refused |

TRUST IN SOURCES OF INFORMATION

52. **How much do you trust information about COVID-19 from the following sources?**

**Public Health Official**

**Very little trust	A great deal of trust	DK**

**0	1	2	3	4	5	6	7	8	9	10	88**

**Doctors**

**Very little trust	A great deal of trust	DK**

**0	1	2	3	4	5	6	7	8	9	10	88**

**Government**

**Very little trust	A great deal of trust	DK**

**0	1	2	3	4	5	6	7	8	9	10	88**

**Employer**

**Very little trust	A great deal of trust	DK**

**0	1	2	3	4	5	6	7	8	9	10	88**

**Union**

**Very little trust	A great deal of trust	DK**

**0	1	2	3	4	5	6	7	8	9	10	88**

**Television**

**Very little trust	A great deal of trust	DK**

**0	1	2	3	4	5	6	7	8	9	10	88**

**Newspapers**

**Very little trust	A great deal of trust	DK**

**0	1	2	3	4	5	6	7	8	9	10	88**

**Social Media**

**Very little trust	A great deal of trust	DK**

**0	1	2	3	4	5	6	7	8	9	10	88**

**Radio**

**Very little trust	A great deal of trust	DK**

**0	1	2	3	4	5	6	7	8	9	10	88**

TRUST IN INSTITUTIONS (PERCEPTIONS)

53. **How much confidence do you have that the following can handle the COVID-19 challenge well?**

**Doctors**

**Very low confidence	Very high confidence	DK/NA**

**0	1	2	3	4	5	6	7	8	9	10	88**

**Government**

**Very low confidence	Very high confidence	DK/NA**

**0	1	2	3	4	5	6	7	8	9	10	88**

**Your employer or workplace**

**Very low confidence	Very high confidence	DK/NA**

**0	1	2	3	4	5	6	7	8	9	10	88**

**Union**

**Very low confidence	Very high confidence	DK/NA**

**0	1	2	3	4	5	6	7	8	9	10	88**

TESTING AND TRACING

54. **If you come in contact with someone who tests positive for COVID-19 but have no symptoms yourself – will you get tested if you have the opportunity?**

O I would get tested for sure

O I may not get tested

For those who select “I would get tested for sure”:

**Please elaborate on this**: *choose as many as apply*

**I would get tested for sure because (check all that apply)**…

O I want to receive the appropriate care in case of a positive test

O This is my responsibility as a citizen

O I would face penalties if I did not

O I believe this helps stop the spread of COVID-19

  O This way I can protect other people

O My friends and family would expect me to get tested

O Other _________________

For those who select “I may not get tested”:

**Please elaborate on this**: *choose as many as apply*

**I may not get tested because (check all that apply) …**

O getting tested would cost money (e.g. transportation, buying the test, taking time off work)

O I do not know where to go to be tested

O it is too time-consuming to get tested

O this will result in loss of income for me due to quarantine while waiting to get the results

O this would result in loss of income for me if I get a positive test

O people might blame me for my actions i f I get a positive test

O I might face fines or other penalties if I had violated official COVID restrictions

O I do not trust authorities with my personal data

O I do not believe COVID-19 exists

O there is nothing I can do, even if I get a positive test

O I am not able to self-isolate in case I get a positive test

O I do not think the tests are reliable

O I am worried people will treat me badly if I get a positive test

O I am worried I will get infected at the testing site

O I think testing will be painful

O Other ____________________

55. **If you test positive for COVID-19 and are asked to share with health authorities the names of people you have been in contact with – will you share all names?**

O I would share all names for sure

O I may not share all names

**I would share all names for sure because…**

O I believe this helps stop spread of COVID-19

O this is my responsibility as a citizen

O this way I can protect other people

O my friends and family would expect me to do this

O I would face penalties if I did not

UNHEALTHY BEHAVIORS

56. **Within the last two weeks, have you done the following:**

**Exercised less than I did before the pandemic** o Yes o No o DK/Refused

**Ate more unhealthy food than I did before the pandemic** o Yes o No o DK/Refused

**Smoked more than I did before the pandemic** o Yes o No o DK/Refused

**Postponed vaccination for myself or my child** o Yes o No o DK/Refused

COVID-19 VACCINE

57. I would get the COVID-19 vaccine when it is available and recommended to me


    IF disagree, or strongly disagree, Can you please explain why you will not to get a COVID-19 vaccine?
    IF unsure, can you explain why are you unsure if you will get a COVID-19 vaccine?

**Strongly Disagree	Strongly Agree	DK/Unsure**

**0	1	2	3	4	5	6	7	8	9	10	88**

58. **My decision of whether or not to get vaccinated would depend on:**

*Choose as many as apply*

| o Whether the vaccine has been in use for a long time with no serious side-effects | Not at all - - - - - - - - - - - - - Very much so - - - DK/Refused |
| --- | --- |
| o Risk of getting infected with COVID-19 at the time when the vaccine is available | Not at all - - - - - - - - - - - - - Very much so - - - DK/Refused |
| o Whether a high vaccination uptake would lift restrictions on movement and gathering in groups | Not at all - - - - - - - - - - - - - Very much so - - - DK/Refused |

**I believe a vaccine can help control the spread of COVID-19**

**Strongly Disagree	Strongly Agree	DK**

**0	1	2	3	4	5	6	7	8	9	10	88**

**If I knew I had been infected with COVID-19 before, I would not get the vaccine even if it were available**

**Strongly Disagree	Strongly Agree	DK**

**0	1	2	3	4	5	6	7	8	9	10	88**

**When everyone else is vaccinated against COVID-19, then I don't have to get vaccinated**

**Strongly Disagree	Strongly Agree	DK**

**0	1	2	3	4	5	6	7	8	9	10	88**

60. **Apart from COVID-19, I think everyone should be vaccinated according to the national vaccination schedule**

o Yes

o No

o Don’t know

o Prefer not to answer

COVID-19 HEALTH CARE/ PUBLIC HEALTH INTERVENTIONS:

61. **How effective do you think were the above intervention in stopping the spread of COVID-19 in Alberta:**

| **Restriction of Mass gatherings** | o Very effective o Somewhat effective o Not effective o DK |
| --- | --- |
| **School closures** | o Very effective o Somewhat effective o Not effective o DK |
| **Physical Distancing** | o Very effective o Somewhat effective o Not effective o DK |
| **Travel Restrictions** | o Very effective o Somewhat effective o Not effective o DK |
| **Wearing a mask in indoor spaces** | o Very effective o Somewhat effective o Not effective o DK |

[HEALTH INSURANCE COVERAGE]

Now, turning to your health insurance coverage. Please include any private, government or employer-paid plans.

62. **Do you have supplemental medical insurance (i.e. coverage to pay for your medications, etc.):**

o Yes

o No

o Don’t know

o Prefer not to answer

[EVERYDAY DISCRIMINATION]

We are interested in learning more about any discrimination you have faced in the past or currently continue to face. With this information, we hope to learn better the hidden struggles of Cargill employees who may face discrimination by the hands of their employers or colleagues.

63. **In your day-to-day life (during COVID-19 pandemic), how often do any of the following happen to you?**

**You are treated with less courtesy than other people are:**

o Almost every day

o At least once a week

o A few times a month

o A few times a year

o Less than once a year

o Never

**You are treated with less respect than other people are:**

o Almost everyday

o At least once a week

o A few times a month

o A few times a year

o Less than once a year

o Never

**You receive poorer service than other people at restaurants or stores:**

o Almost every day

o At least once a week

o A few times a month

o A few times a year

o Less than once a year

o Never

**People act as they think you are not smart:**

o Almost every day

o At least once a week

o A few times a month

o A few times a year

o Less than once a year

o Never

**People act as if they are afraid/scared of you:**

o Almost every day

o At least once a week

o A few times a month

o A few times a year

o Less than once a year

o Never

**People act as if they think you are dishonest:**

o Almost everyday

o At least once a week

o A few times a month

o A few times a year

o Less than once a year

o Never

**People act as if they are better than you are:** o Almost everyday

o At least once a week

o A few times a month

o A few times a year

o Less than once a year

o Never

**You are called names or insulted:**

o Almost everyday

o At least once a week

o A few times a month

o A few times a year

o Less than once a year

o Never

**You are threatened or harassed:**

o Almost everyday

o At least once a week

o A few times a month

o A few times a year

o Less than once a year

o Never

EVERYDAY DISCRIMINATION

**FOLLOW-UP QUESTION: Asked only of those answering “A few times a year” or more frequently to at least ONE question.**

***What do you think is the main reason for these experiences? (check more than one if volunteered)***

| o Ancestry or Origins or Tribe | o Religion | o Sexual Orientation |
| --- | --- | --- |
| o Gender | oHeight | oEducation or Income Level |
| o Race | o Weight | oPhysical disability |
| o Age | o Other aspect of physical appearance | o Skin color |

🞏 Other (Specify) _____________________________________

[EMPLOYMENT HAZARDS]

64. **Using the following scale, please answer the following questions to the best of your ability regarding your perceptions of the demand of your current employment. Would you say…..**

|  | *Strongly Disagree* | *Disagree* | *Neither Agree nor Disagree* | *Agree* | *Strong Agree* | *DK/Unsure* |
| --- | --- | --- | --- | --- | --- | --- |
| The job is stressful | 1 | 2 | 3 | 4 | 5 | 88 |
| Fast-paced work | 1 | 2 | 3 | 4 | 5 | 88 |
| The work is hard | 1 | 2 | 3 | 4 | 5 | 88 |
| Work is excessive | 1 | 2 | 3 | 4 | 5 | 88 |
| There is enough time to do the work | 1 | 2 | 3 | 4 | 5 | 88 |

PERCIEVED HIGH JOB INSECURITY:

65. **Using the following scale, please answer the following questions to the best of your ability regarding your perceptions of the job security of your current employment. Would you say…..**

|  | *Strongly Disagree* | *Disagree* | *Neither Agree nor Disagree* | *Agree* | *Strong Agree* | *DK/Unsure* |
| --- | --- | --- | --- | --- | --- | --- |
| You worry about the future in your job | 1 | 2 | 3 | 4 | 5 | 88 |
| You feel secured in your employment | 1 | 2 | 3 | 4 | 5 | 88 |
| You think the company will be here in 5 – 10 years | 1 | 2 | 3 | 4 | 5 | 88 |

PERCIEVED HIGH VULNERABILITY AT WORK:

66. **Using the following scale, please answer the following questions to the best of your ability regarding your perceptions of how vulnerable you feel at your current employment. Would you say you are…..**

|  | *Never* | *Rarely* | *Sometimes* | *Often* | *Always* | *DK/Unsure* |
| --- | --- | --- | --- | --- | --- | --- |
| Made to feel you could be easily replaced by your boss | 1 | 2 | 3 | 4 | 5 | 88 |
| Afraid of being fired even though you did nothing wrong | 1 | 2 | 3 | 4 | 5 | 88 |
| Treated in discriminatory or unjust ways on your job | 1 | 2 | 3 | 4 | 5 | 88 |
| Concerned about your safety but afraid to speak up | 1 | 2 | 3 | 4 | 5 | 88 |
| Felt defenseless against unfair treatment directed toward you | 1 | 2 | 3 | 4 | 5 | 88 |

[ECONOMIC DISPARITY]

The next set of questions is regarding your household income, individuals in your household and their health insurance status.

67. **Please assess your private financial situation over the past three months:**

o Improved

o Remains the same

o Worse

o Don’t know/ Refused

68. **Did you have ever have difficulty making ends meet at the end of the month?**

O Frequently

O Sometimes

O Never

O Prefer not to answer.

69. **What is your household income range:**

o Less than $10,000

o Less than $25,000

o Less than $40,000

o Less than $50,000

o Less than $75,000

o Less than $100,000

o More than $100,000

- Don't know
- Refuse to answer

70. **What do you think can be done or should be done to make your workplace safer from COVID-19? (for example, to prevent infections or an outbreak)
    ______________________________________**
71. **What do you think can or should be done to improve your work conditions or make your workplace better?
    _______________________________________**

**Was there anything else you think we missed that you think we should know? _____________________________**

**Thank you for your time and participation. !!**

**Supplement 2.2: Interview Guide**

*Semi-structured interview guide used for individual and family interviews.*

**Individual / Family Interview Guide**

**ID #: __ __ __ __ __ __ __ __**

**Date of Interview: __ __/__ __/ __ __ __ __**

DD	MM	YEAR

**Interviewer initials: ______________**

**Medium (Select ONE):**

o In-person (recording/transcription - Otter)

o Video conferencing (recording/transcription - Otter)

o Phone call (recording/transcription - Otter)

**Duration of Interview: _____** minutes

**This interview is with:**

o Meat processing plant employee

o Family member(s) of employee (please specify relationship:___________________)

______________________________________________________________________________Which meat processing plant does the participant, or family member work at?

o Cargill

o JBS

O Harmony

O Other _________________________

**Notes:**

*[If using an interpreter, allow interpreter time to introduce themselves and their role].*

Hello,

My name is __________. I am (a student, community scholar/member, research assistant) working on a study being conducted by the University of Calgary’s Cumming School of Medicine. We are keen to hear your story, your experience, around the recent COVID-19 outbreak in your community/ meat processing plant.

*GO THROUGH CONSENT FORM*

*IF VERBAL CONSENT RECEIVED, CONTINUE.*

You do not have to answer any questions that you do not want to. You can also stop the interview if you want at any time. Your thoughts, feelings and stories around your experience with the COVID-19 outbreak are important to us.

If you have any questions or concerns, please let me know. If not, let’s get started.

**Note to research team:** Interviews will explore important thoughts, feelings and stories related to participants experience during the outbreak. Trust will play a major role in achieving a meaningful and authentic interview.

**Interview Guide**:

1. Please tell me what makes you unique, what makes you special?

Potential probes:

a. How long have you lived here in Alberta?
b. Where are you from if you haven’t lived here your entire life?
c. Who lives with you here in AB?
2. Please describe what your job is like? What is a typical workday for you?

Potential probes:

a. Where do you work? What is your position? What do you do?
b. How long have you worked there for?
3. Are you able to find COVID-19 health information? If yes, where?

Potential probes:

a. Who do you trust to provide reliable information and advice about COVID-19? For example, through: the news, your work, your doctor’s office, the government, friends and family, social media, radio?
b. How has your language skills impaired or improved you access to public health information? Other barriers?

4. Please share your experience during the COVID-19 outbreak?

Potential probes:

a. What barriers or facilitators do you encounter during the outbreak?
b. How has COVID-19 affected your job/work?
c. How has COVID-19 affected your family?
d. Did you or anyone close to you get sick?
5. How do you describe what happened at work during the COVID-19 outbreak?

Potential probes:

a. How did you feel about the safety measures put in place at your work?
b. How did your employer communicate with you (e.g., email, text messaging, meetings, individually) and what was communicated to you?
c. Did your employer involve you and your co-workers in the response? Would you have liked to be involved?

6. Did your employer support testing and hep link you to a family doctor?

*[if they say Yes – have they gotten sick from COVID-19?]*

Tell me about your experience having COVID-19?

Potential probes:

a. Did public health contact you? What supports or advice did they provide?

7. Did you or someone close to you have to be quarantined or have to self-isolate?

*[If yes] How was it for you/them being quarantined?*

Potential probes:

a. Did you have a safe space to self-isolate without risk of infecting other members of your family/household?
b. What do you think could made it better or easier for employees and their households to quarantine or self-isolate?
c. If you had to be isolated or were infected did you feel any stigma or negative emotions from anyone when you returned to work?

8. Did you and your co-workers feel safe?

Potential probes:

a. Did you and your co-workers feel like you had a choice about returning to work or not?

9. How do you feel healthcare providers responded to the outbreak?

Potential probes:

a. Did healthcare providers communicate with you? (e.g., email, text messaging, meetings, individually), and what was communicated to you?
b. What actions did they take? How did you personally feel about their response?
c. Did healthcare providers involve you and your co-workers in the response? Would you have liked to be involved?
d. What do you think could improve how healthcare providers respond to any future outbreaks at meat processing plants?

10. How do you think the union (UFCW401 or other) responded to the COVID-19 outbreak?

Potential probes:

a. How did your union communicate with you? (e.g., email, text messaging, meetings, individually) and what was communicated to you?
b. How did the union involve you and your co-workers in the response? Were there other responses you wanted?
c. Did other community agencies or organizations communicate with you? (i.e. CCIS, Action Dignity, PCNs etc.)? If so, do you know who?

11. Who or what do you think was most helpful in the outbreak response? (i.e., AHS, family MD, employer, union, social agency etc.)

Potential probes:

a. What made them most helpful?

12. What responses created barriers or challenges during the outbreak response?

Potential probes:

a. What made them unhelpful?
b. What should have been done differently?

13. Is there anything else that you would like to share?

**Demographic Questions:**

1. What gender would you identify as?
2. What year were you born in?
3. What ethnicity would you identify as? (Asian/Indigenous/Latin American/Middle Eastern/White/Black/African)
4. What is the primary language that you speak?
5. What is your immigration status in Canada?

**Supplement 2.3: Survey Domains and References**

*Source instruments and validated scales used to construct the survey domains.*

| Domain | Source | Description & Further citations (if applicable) |
| --- | --- | --- |
| Sociodemographic questions, & other social determinants of health | [Canadian Community Health Survey - Annual Component, Statistics Canada, 2020](https://www23.statcan.gc.ca/imdb/p3Instr.pl?Function=assembleInstr&lang=en&Item_Id=1262397)  [COVID-19 Case Report Form](https://www.canada.ca/content/dam/phac-aspc/documents/services/diseases/2019-novel-coronavirus-infection/health-professionals/2019-nCoV-case-report-form-en.pdf) | 1. Demographics (sex, gender, age, proxies for minors/elders/others, age, occupation) 2. Chronic Conditions 3. Main Activity (MAC) 4. General Health: Self-reported health and mental health, stress, smoking 5. Self-reported English and language proficiency 6. Income: What is your household income range? 7. Sense of belonging 8. Workplace stress pre/during Covid-19 9. Health insurance coverage |
| Family/Household information | [WHO Covid-19 Survey Tool and Guidance, WHO, 2020](https://www.who.int/europe/publications/i/item/WHO-EURO-2020-696-40431-54222) | # people in household  Seniors  Minors |
| Everyday Discrimination | [Everyday Discrimination Scale](https://hsph.harvard.edu/research/williams-group/measuring-discrimination-resource/) (EDS) | The EDS is used to measure the unfair treatment that individuals experience routinely during their day-to-day life. We will use selected questions to ask how participants perceive they are treated generally in their daily life.  Used as a measure of subjective experiences of daily discrimination |
| Employment Hazards: | [The Employment Precariousness Scale (EPRES), Vives et al., 2010](https://oem.bmj.com/content/67/8/548)  [Gosselin et al., 2020](https://onlinelibrary.wiley.com/doi/abs/10.1002/ajim.23146)  [Jonsson et al., 2019](https://bmjopen.bmj.com/content/9/9/e029577)  [Padrosa et al., 2020](https://link.springer.com/article/10.1007/s11135-020-01017-2) | Stress in the Workplace (Job Strain)  Perceived High Job Insecurity  Perceived High Vulnerability at Work |
| Poverty Screener | [Brcic, Eberdt, and Kaczorowski, 2011](https://www.ncbi.nlm.nih.gov/pmc/articles/PMC3268233/) | Did you have ever have difficulty making ends meet at the end of the month? |
| Finance Self-Assessment | [WHO Covid-19 Survey Tool and Guidance, WHO, 2020](https://www.who.int/europe/publications/i/item/WHO-EURO-2020-696-40431-54222) | Please assess your private financial situation over the past three months: |
| COVID-19 Questions | | |
| COVID-19 Personal Experience | [WHO Covid-19 Survey Tool and Guidance, WHO, 2020](https://www.who.int/europe/publications/i/item/WHO-EURO-2020-696-40431-54222) | Additional sources cited from WHO Guidance document - Validated items adapted from:  Brewer, N. T., Chapman, G. B., Gibbons, F. X., Gerrard, M., McCaul, K. D., & Weinstein, N. D. (2007). Meta-analysis of the relationship between risk perception and health behavior: the example of vaccination. Health psychology, 26(2), 136. |
| Health Literacy |  | Additional sources cited from WHO Guidance document - Validated items adapted from:  Sørensen K, Van den Broucke S, Pelikan JM, et al. Measuring health literacy in populations: illuminating the design and development process of the European Health Literacy Survey Questionnaire (HLS-EUQ). BMC Public Health. 2013;13:948. Published 2013 Oct 10. doi:10.1186/1471- 2458-13-948  Griebler, Robert; Nitsche, Michael (2020): The Austrian Corona Health Literacy Questionnaire. Vienna: Gesundheit Österreich GmbH & Das Österreichische Gallup Institut |
| Preparedness and Perceived Self-Efficacy |  | Additional sources cited from WHO Guidance document - Validated items adapted from:  Bandura, A. (2006). Guide for constructing self-efficacy scales. Self-efficacy beliefs of adolescents, 5(1), 307-337. Psychological construct: perceived self-efficacy  Renner, B., & Schwarzer, R. (2005). The motivation to eat a healthy diet: How intenders and nonintenders differ in terms of risk perception, outcome expectancies, selfefficacy, and nutrition behavior. Polish Psychological Bulletin, 36(1), 7-15. |
| Prevention - Own Behaviours |  | Additional sources cited from WHO Guidance document - Items adapted from:  Steel Fisher GK et al (2012). Public response to the 2009 influenza A H1N1 pandemic: a polling study in five countries. Lancet Infectious Diseases 2012; 12: 845–50 |
| Trust in Sources of Information |  | Additional sources cited from WHO Guidance document:  Schweitzer, M. E., Hershey, J. C., & Bradlow, E. T. (2006). Promises and lies: Restoring violated trust.Organizational behavior and human decision processes, 101(1), 1-19.  Pearson, S. D., & Raeke, L. H. (2000). Patients' trust in physicians: many theories, few measures, and little data.Journal of general internal medicine, 15(7), 509- 513. |
| Trust in Institutions |  | Additional sources cited from WHO Guidance document:  Schweitzer, M. E., Hershey, J. C., & Bradlow, E. T. (2006). Promises and lies: Restoring violated trust.Organizational behavior and human decision processes, 101(1), 1-19.  Pearson, S. D., & Raeke, L. H. (2000). Patients' trust in physicians: many theories, few measures, and little data.Journal of general internal medicine, 15(7), 509- 513. |
| Testing and Tracing |  | Additional sources cited from WHO Guidance document:  Michie et al (2014), The Behaviour Change Wheel. A Guide to Designing Interventions. Silverback Publishing. ISBN 978-1-912141- 00-5 |
| Unhealthy Behaviours |  | Unwanted behaviours; Allows to identify adverse behaviours that may need to be addressed. |
| COVID-19 Vaccine (Vaccine Hesitancy) |  | Additional sources cited from WHO Guidance document:    Michie et al (2014), The Behaviour Change Wheel. A Guide to Designing Interventions. Silverback Publishing. ISBN 978-1-912141- 00-5  Betsch, C., Schmid, P., Heinemeier, D., Korn, L., Holtmann, C., & Böhm, R. (2018). Beyond confidence: Development of a measure assessing the 5C psychological antecedents of vaccination. PLOS ONE, 13(12), e0208601. <https://journals.plos.org/plosone/article?id=10.1371/journal.pone.0208601>  Open Data Open Materials |
