## Supplement 3 for "Occupational hierarchy, racialization, and COVID-19 health outcomes among meat processing plant workers in Alberta: a community-engaged mixed-methods study"

**Supplement 3: Supplementary Results**

**Supplement 3.1: Themes Identified Using PROGRESS-Plus Factors**

*Qualitative themes mapped to PROGRESS-Plus equity factors with illustrative quotes.*

| Progress Plus Factor | Themes | Quotes |
| --- | --- | --- |
| Occupation | Unsafe working conditions | “*It was that way, but let me tell you, in those days when a lot of people started to get sick. They put in more money per hour. I mean, they, if we usually make $20, well, they were paying $24 an hour. That is, 4 more dollars an hour to people who went t*  *o work. So, this, I feel like it was bad, because a lot of people. I knew of several. They were positive and they didn't care. I mean, they didn't care. And to get there to earn those four more dollars an hour, they would get to work being sick. So they went to work sick and they would go and get those people who were healthy there, go and get them sick, do I explain myself? Because, although there were measures, they had the mask and everything. Well, it's impossible. I mean. I feel like that thanks to the decision they made to pay them for some compensation, but I don't feel like that it was compensation, I feel that it was so people would not stop going to work, so that they wouldn't stop producing*.” Participant 5    “*...because I didn't want to get infected again because I knew that this sick people were still going to work. But I also took my personal measures, I mean, as a personal responsibility...*” Participant 5 |
| Ethnicity | Discrimination based on the nationality | “*I had seen a lot of bad treatments. I had seen females from specific countries crying because of bad treatment by the lead hands and supervisors. At <MEAT PLANT 1> if there is a Sudanese supervisor Sudanese are treated well, if you have a Philippines supervisor Philippians are treated well and do easy jobs.*” Participant 16 |
| Gender | Gender based discrimination | “*But, when I returned from maternity leave, I don't know what would have happened or I don't know if I was working too hard, so I had a problem in my hand, I mean, I got a bone on top of my bone, here (she shows her wrist to the camera)... I didn't want to get up anymore, because I felt tired, I felt the harassment, he wouldn't even let me go to the bathroom, and if went to the bathroom, I mean, he would let my work pile up there, he would do this only to me. I mean, there were a lot of things that started to get happen all together. So, I had to go to another human resources area, because I had already gone to human resources to report him many times, but no one listened to me.*” Participant 5 |
| Language | Discrimination based on the language | “*I saw so many women crying because they could not express what is happening to them due to language barrier. It is very difficult to work there especially if you don’t have a lead hand or a supervisor from your country of origin or tribe.”* Participant 16 |
| Health | COVID-19 infection due to lack of physical and mental health | “*Yes, I was infected with* *COVID. At MEAT PLANT 2 there were so many employees who were infected with COVID and there was* *a number of people close to who were infected.*” Participant 4    “*I mean, I felt lucky that although it had been hard, even though it had been heavy, even though it had been, well, something I had never experienced, I felt lucky because many, many people that I knew had to go to the hospital because they couldn't breathe or well they just got infected and didn't live it to tell about it, so I felt very lucky.*” Participant 5 |
| Socioeconomic Status | Income precarity due to economic vulnerability | “*I think if you live paycheck by paycheck, and that my understanding [NAME REMOVED] was only one working, that $500 goes a long way. And people are going to come to work sick, because hey, if I didn't come to work, I didn't get the $500 bonus.* *So he had to come to work. And eventually he got sick, and he passed* *away.*“ Participant 14 |
| Social Capital | Lack of trust on safety measures      Impact on family relationships due to Covid-19 pandemic | “*You still never feel safe. You will never feel safe anywhere.* *So you still, like you can see* *them they're doing* *their, their diligence and like sanitizing, but you don't know what's, what's going to happen.* *Right? You don't know if the person* *next to you. sitting right next to you is infected or you don't know, you always have this anxiety*.” Participant 25    “*I was very careful before and after I caught COVID. COVID has left a permanent scar among our family circles. Particularly, it intervened on the love, affection and attachment between me and my lovely youngest kid, who avoided me a lot, especially after my family heard about MEAT PLANT 2’s COVID outbreak.*” Participant 20 |
| Plus Factors | Treatment and power hierarchy in large corporations | “*There were cases whereas* *new worker you will be treated like a baby who does not have rights and duties. Thus, there were differential treatments for beginners and senior workers. The managers treat seniors with better respect, sensitivity, and dignity.*” Participant 33  “*I got hurt, so I had to go to get checked, and when I went to get checked my supervisor got more upset, I mean, I felt much more workplace harassment from him when I got hurt*.” Participant 5 |

**Supplement 3.2: Unadjusted Outcomes by Occupational Group (n=187)**

| **Characteristic** | **General Labour n=123** | **Skilled Labour n=51** | **Management n=13** | **Total** | **p-value** |
| --- | --- | --- | --- | --- | --- |
| **COVID-19 Diagnosis** | | | | | |
| Positive, n (%) | 43 (35.5%) | 11 (21.6%) | 1 (7.7%) | 55 (29.7%) | **0.037*** |
| n with outcome data | 121 | 51 | 13 | 185 |  |
| **Fair or Poor Self-Rated Health** | | | | | |
| Fair/Poor, n (%) | 22 (17.9%) | 5 (10.0%) | 1 (7.7%) | 28 (15.1%) | 0.313 |
| Good or better, n (%) | 101 (82.1%) | 45 (90.0%) | 12 (92.3%) | 158 (84.9%) |  |
| n with outcome data | 123 | 50 | 13 | 186 |  |
| **Fair or Poor Self-Rated Mental Health** | | | | | |
| Fair/Poor, n (%) | 29 (24.4%) | 12 (24.0%) | 2 (15.4%) | 43 (23.6%) | 0.767 |
| Good or better, n (%) | 90 (75.6%) | 38 (76.0%) | 11 (84.6%) | 139 (76.4%) |  |
| n with outcome data | 119 | 50 | 13 | 182 |  |
| **Mean Everyday Discrimination Score (range 1-6)** | | | | | |
| Mean [IQR] | 2.30 [1.00-3.25] | 2.10 [1.00-2.89] | 2.48 [1.53-3.22] | 2.26 [1.00-3.11] | 0.492 |
| n | 123 | 51 | 13 | 187 |  |
| *All analyses use the full analytic sample (n=187, post occupational group filter). Outcome-specific ns vary due to missing outcome data.* | | | | | |
| *Chi-square tests used for COVID-19, Fair/Poor Health, and Fair/Poor Mental Health. Kruskal-Wallis test used for mean EDS score.* | | | | | |
| ** p < 0.05. EDS = Everyday Discrimination Scale; mean score computed across 9 items (range 1-6, higher = more frequent discrimination). IQR = interquartile range.* | | | | | |

**Supplement 3.3: Unadjusted Distribution of the Everyday Discrimination Scale Across Occupation Group**

*Item-level Everyday Discrimination Scale results by occupational group.*

| **Everyday Discrimination** | Labor Median [IQR] | Skilled Labor Median [IQR] | Management Median [IQR] | **Total Median [IQR]** | **P-Value of Kruskal Wallace** |
| --- | --- | --- | --- | --- | --- |
| You are treated with less courtesy than other people are | 2 (1-4) | 2 (1-3) | 3 (2-4.5) | 2 (1-3.5) | 0.210 |
| You are treated with less respect than other people are | 3 (1-4) | 1 (1-3) | 3 (2.5-4) | 3 (1-4) | 0.151 |
| You receive poorer service than other people at restaurants or stores | 1 (1-3) | 1 (1-3) | 2.5 (1-3.25) | 1 (1-3) | 0.269 |
| People act as they think you are not smart | 2 (1-4) | 1 (1-3) | 2 (1-4) | 2 (1-4) | 0.176 |
| People act as if they are afraid/scared of you | 1 (1-3) | 1 (1-3) | 3 (1-3) | 1 (1-3) | 0.447 |
| People act as if they think you are dishonest | 1 (1-3) | 1 (1-3) | 1 (1-3.5) | 1 (1-3) | 0.901 |
| People act as if they are better than you are | 3 (1-4) | 2 (1-3) | 2.5 (1-3) | 2 (1-4) | 0.268 |
| You are called names or insulted | 1 (1-2) | 1 (1-3) | 1 (1-3) | 1 (1-3) | 0.721 |
| You are threatened or harassed | 1 (1-2) | 1 (1-3) | 1 (1-3) | 1 (1-3) | 0.595 |
| Aggregate Median Discrimination | 2 (1-3) | 1 (1-3) | 2.5 (1.3-3.125) | 1.5 (1-3) | 0.523 |

*Bold indicates statistically significant two-tailed p-values (α = 0.05).*

*IQR = interquartile range*

**Supplement 3.4: Multivariable Logistic Regression Results — All Primary Models**

*Adjusted odds ratios for all four primary outcome models.*

| **Predictor** | **COVID-19 Diagnosis (n=168)** | | | **Fair/Poor Health (n=169)** | | | **Fair/Poor Mental Health (n=166)** | | | **Everyday Discrimination (Mean EDS Score) (n=170)** | | |
| --- | --- | --- | --- | --- | --- | --- | --- | --- | --- | --- | --- | --- |
|  | **aOR** | **95% CI** | **p-value** | **aOR** | **95% CI** | **p-value** | **aOR** | **95% CI** | **p-value** | **β** | **95% CI** | **p-value** |
| **Occupational Group** | | | | | | | | | | | | |
| Skilled Labour | **0.38** | **0.15-0.89** | **0.032** | 0.54 | 0.16-1.55 | 0.280 | 0.83 | 0.33-2.00 | 0.692 | -0.358 | -0.796-0.080 | 0.109 |
| Management | 0.13 | 0.01-0.75 | 0.060 | 0.57 | 0.03-3.74 | 0.621 | 0.74 | 0.10-3.43 | 0.727 | 0.083 | -0.659-0.825 | 0.825 |
| **Racialized Identity** | | | | | | | | | | | | |
| Other Racialized | 0.61 | 0.28-1.31 | 0.213 | **0.20** | **0.06-0.54** | **0.003** | **0.24** | **0.09-0.58** | **0.002** | -0.081 | -0.497-0.336 | 0.702 |
| White | 0.82 | 0.23-2.58 | 0.738 | 0.44 | 0.09-1.60 | 0.250 | 0.76 | 0.25-2.14 | 0.608 | 0.136 | -0.441-0.714 | 0.641 |
| **Facility** | | | | | | | | | | | | |
| Non-MPC-1 | **0.29** | **0.12-0.67** | **0.005** | 0.78 | 0.29-1.98 | 0.612 | 0.92 | 0.40-2.06 | 0.846 | **-0.538** | **-0.955--0.121** | **0.012** |
| *Reference groups: General Labour (occupational group), Black (racialized identity), MPC-1 (facility).* | | | | | | | | | | | | |
| *Dark grey (bold): p < 0.05.* | | | | | | | | | | | | |
| *COVID-19 Diagnosis, Fair/Poor Health, Fair/Poor Mental Health: multivariable logistic regression; estimates are adjusted odds ratios (aOR) with Wald 95% CIs.* | | | | | | | | | | | | |
| *Everyday Discrimination: linear regression of mean EDS score (range 1-6, higher = more frequent discrimination); estimates are unstandardised beta coefficients (β) with 95% CIs. R² = 0.055.* | | | | | | | | | | | | |
| *Income precarity excluded from primary models due to 17.6% missingness; see Supplement 9.* | | | | | | | | | | | | |
| *Management estimates (n=13) should be interpreted as directional only given small subgroup size.* | | | | | | | | | | | | |

**Supplement 3.5: Sex-Stratified Multivariable Logistic Regression Results (SAGER Analyses)**

*Sex-stratified models in line with the SAGER guidelines.*

| **Predictor** | **Male (n=130)** | | | | | | | | | | | | **Female (n=38)** | | | | | | | | | | | |
| --- | --- | --- | --- | --- | --- | --- | --- | --- | --- | --- | --- | --- | --- | --- | --- | --- | --- | --- | --- | --- | --- | --- | --- | --- |
|  | **COVID-19 Diagnosis** | | | **Fair/Poor Health** | | | **Fair/Poor Mental Health** | | | **Everyday Discrimination** | | | **COVID-19 Diagnosis** | | | **Fair/Poor Health** | | | **Fair/Poor Mental Health** | | | **Everyday Discrimination** | | |
|  | **OR** | **95% CI** | **p** | **OR** | **95% CI** | **p** | **OR** | **95% CI** | **p** | **OR** | **95% CI** | **p** | **OR** | **95% CI** | **p** | **OR** | **95% CI** | **p** | **OR** | **95% CI** | **p** | **OR** | **95% CI** | **p** |
| **Occupational Group** | | | | | | | | | | | | | | | | | | | | | | | | |
| Skilled Labour | **0.32** | **0.11-0.85** | **0.028** | 0.59 | 0.14-2.17 | 0.450 | 0.56 | 0.18-1.59 | 0.294 | **0.22** | **0.04-0.82** | **0.042** | 0.48 | 0.02-4.09 | 0.549 | 0.69 | 0.03-6.57 | 0.769 | 1.20 | 0.05-12.70 | 0.886 | **—** | **—** | **—** |
| Management | 0.12 | 0.01-0.78 | 0.063 | 0.90 | 0.04-9.21 | 0.932 | 0.19 | 0.01-1.48 | 0.172 | 0.17 | 0.01-1.20 | 0.134 | **—** | **—** | **—** | **—** | **—** | **—** | **—** | **—** | **—** | **—** | **—** | **—** |
| **Racialized** | | | | | | | | | | | | | | | | | | | | | | | | |
| Other Racialized | 0.51 | 0.20-1.22 | 0.134 | **0.06** | **0.00-0.32** | **0.008** | **0.18** | **0.05-0.52** | **0.004** | 1.48 | 0.49-4.53 | 0.487 | 1.12 | 0.15-10.04 | 0.914 | 0.29 | 0.04-1.89 | 0.200 | 0.31 | 0.04-2.10 | 0.225 | 0.52 | 0.03-13.97 | 0.646 |
| White | 0.91 | 0.17-4.22 | 0.902 | **—** | **—** | **—** | 1.93 | 0.48-7.91 | 0.350 | **6.41** | **1.16-39.91** | **0.034** | 1.00 | 0.09-10.76 | 1.000 | 0.15 | 0.01-1.44 | 0.138 | 0.15 | 0.01-1.42 | 0.136 | 3.13 | 0.16-133.90 | 0.471 |
| **Facility** | | | | | | | | | | | | | | | | | | | | | | | | |
| Non-MPC-1 | **0.26** | **0.08-0.70** | **0.012** | 0.71 | 0.20-2.26 | 0.573 | 0.69 | 0.25-1.78 | 0.452 | 0.47 | 0.12-1.53 | 0.239 | 0.38 | 0.06-2.02 | 0.274 | 0.70 | 0.10-4.05 | 0.696 | 2.14 | 0.33-14.78 | 0.416 | 0.07 | 0.00-0.76 | 0.057 |
| *Ref: General Labour, Black, MPC-1. Income precarity excluded from primary models. Dark grey (bold): p<0.05. Medium grey (—): complete separation.* | | | | | | | | | | | | | | | | | | | | | | | | |
| *Female stratum (n=38): interpret with caution; multiple cells show complete separation. Confidence intervals are Wald 95% CIs.* | | | | | | | | | | | | | | | | | | | | | | | | |

**Supplement 3.6: Correlation Matrix — Model Predictors and Outcomes**

*Pairwise correlations among predictors and outcomes used in the regression models.*

|  | **Occupational Group** | **Racialized Identity** | **Facility (binary)** | **Facility (full)** | **Sex** | **Income Precarity*** | **COVID-19 Diagnosis** | **Fair/Poor Health** | **Fair/Poor Mental Health** |
| --- | --- | --- | --- | --- | --- | --- | --- | --- | --- |
| **Occupational Group** | **—** | **0.267** | -0.124 | **-0.206** | **0.209** | -0.055 | **-0.214** | -0.106 | -0.063 |
| **Racialized Identity** | **0.267** | **—** | 0.009 | -0.004 | -0.192 | **0.203** | -0.125 | -0.198 | -0.142 |
| **Facility (binary)** | -0.124 | 0.009 | **—** | **0.888** | -0.146 | 0.004 | -0.188 | 0.021 | 0.038 |
| **Facility (full)** | **-0.206** | -0.004 | **0.888** | **—** | -0.139 | 0.025 | -0.085 | 0.115 | 0.075 |
| **Sex** | **0.209** | -0.192 | -0.146 | -0.139 | **—** | **-0.211** | 0.038 | -0.121 | -0.001 |
| **Income Precarity*** | -0.055 | **0.203** | 0.004 | 0.025 | **-0.211** | **—** | -0.020 | 0.147 | 0.125 |
| **COVID-19 Diagnosis** | **-0.214** | -0.125 | -0.188 | -0.085 | 0.038 | -0.020 | **—** | **0.206** | 0.176 |
| **Fair/Poor Health** | -0.106 | -0.198 | 0.021 | 0.115 | -0.121 | 0.147 | **0.206** | **—** | **0.542** |
| **Fair/Poor Mental Health** | -0.063 | -0.142 | 0.038 | 0.075 | -0.001 | 0.125 | 0.176 | **0.542** | **—** |
| *Correlations calculated using pairwise complete observations (n=168). Categorical variables numerically encoded; interpret as indicative of association strength only.* | | | | | | | | | |
| *Medium grey (bold): \|r\| >= 0.20 — moderate association. Dark grey (bold): \|r\| >= 0.70 — high collinearity.* | | | | | | | | | |
| *Facility (binary) and Facility (full) r=0.888, confirming binary facility adequately captures full facility variation.* | | | | | | | | | |
| *Thick borders separate predictor variables (white/grey rows) from outcome variables (light grey rows/columns).* | | | | | | | | | |
| **Income Precarity excluded from primary models due to 17.6% missingness. A separate paper examines income precarity in this cohort. See Supplement 9.* | | | | | | | | | |
| *Sex excluded from primary models due to small female subsample (n=38) and moderate collinearity with occupational group (r=0.21, Cramer's V=0.22). VIF for sex=1.15. See Supplement 10.* | | | | | | | | | |
